# Sleep pressure propels cerebrovascular oscillations while sleep intensity correlates with respiration- and cardiac-driven brain pulsations

**DOI:** 10.1101/2024.10.16.24315580

**Authors:** Sara Marie Ulv Larsen, Sebastian Camillo Holst, Anders Stevnhoved Olsen, Brice Ozenne, Dorte Bonde Zilstorff, Kristoffer Brendstrup-Brix, Pia Weikop, Simone Pleinert, Vesa Kiviniemi, Poul Jørgen Jennum, Maiken Nedergaard, Gitte Moos Knudsen

**Affiliations:** Neurobiology Research Unit, Copenhagen University Hospital, Rigshospitalet; Copenhagen, Denmark; Faculty of Health and Medical Sciences (SUND), University of Copenhagen; Copenhagen, Denmark; Roche Pharma Research and Early Development, Neuroscience and Rare Diseases, Roche Innovation Center Basel; Basel, Switzerland; Department of Applied Mathematics and Computer Science, Technical University of Denmark; Kgs. Lyngby, Denmark; Department of Public Health, Section of Biostatistics, University of Copenhagen; Copenhagen, Denmark; Center for Translational Neuromedicine, University of Copenhagen; Copenhagen, Denmark; Oulu Functional Neuroimaging (OFNI), Department of Diagnostic Radiology, Oulu University Hospital; Oulu, Finland; Biocenter Oulu, Oulu University; Oulu, Finland; Danish Center for Sleep Medicine, Department of Clinical Neurophysiology, Copenhagen University Hospital, Rigshospitalet; Glostrup, Denmark; Department of Clinical Medicine, University of Copenhagen; Copenhagen, Denmark; Center for Translational Neuromedicine, Department of Neurosurgery, University of Rochester Medical Center; Rochester, NY, USA

**Keywords:** Magnetic Resonance Encephalography, Physiological brain oscillations, Brain-fluid dynamics, Glymphatic system, Sleep pressure, NREM Sleep, Slow waves, EEG delta power, Adrenergic antagonism

## Abstract

The waste-clearing flow of cerebrospinal fluid through the brain is driven by cerebrovascular, respiratory, and cardiac forces and is thought to be regulated by homeostatic sleep mechanisms. In a circadian-controlled sleep and sleep deprivation study, we investigated how heightened sleep pressure and slow-wave-rich sleep affect physiological brain oscillations in humans, as measured with accelerated neuroimaging. Our study also included a randomised, double-blind, placebo-controlled, cross-over administration of the adrenergic antagonist carvedilol, which allowed us to examine the effect of modulating cerebrovascular pulsatility. We report that sleep deprivation increases low frequency brain oscillations (LFOs) more than sleep does, with LFOs during sleep correlating with cognitive measures of sleep pressure. Conversely, slow-wave-rich NREM sleep (stages N2 and N3) enhances respiration- and cardiac-driven brain pulsations – particularly within grey and white matter. The strength of these brain pulsations escalates with sleep depth (N3 > N2) and correlates with EEG delta power. Carvedilol dampens LFOs, supporting that these reflect cerebrovascular oscillations. Altogether, our findings indicate that sleep pressure promotes cerebrovascular oscillations, while sleep slow waves synchronise with respiration- and cardiac-driven brain pulsations.

**Graphical Abstract:** 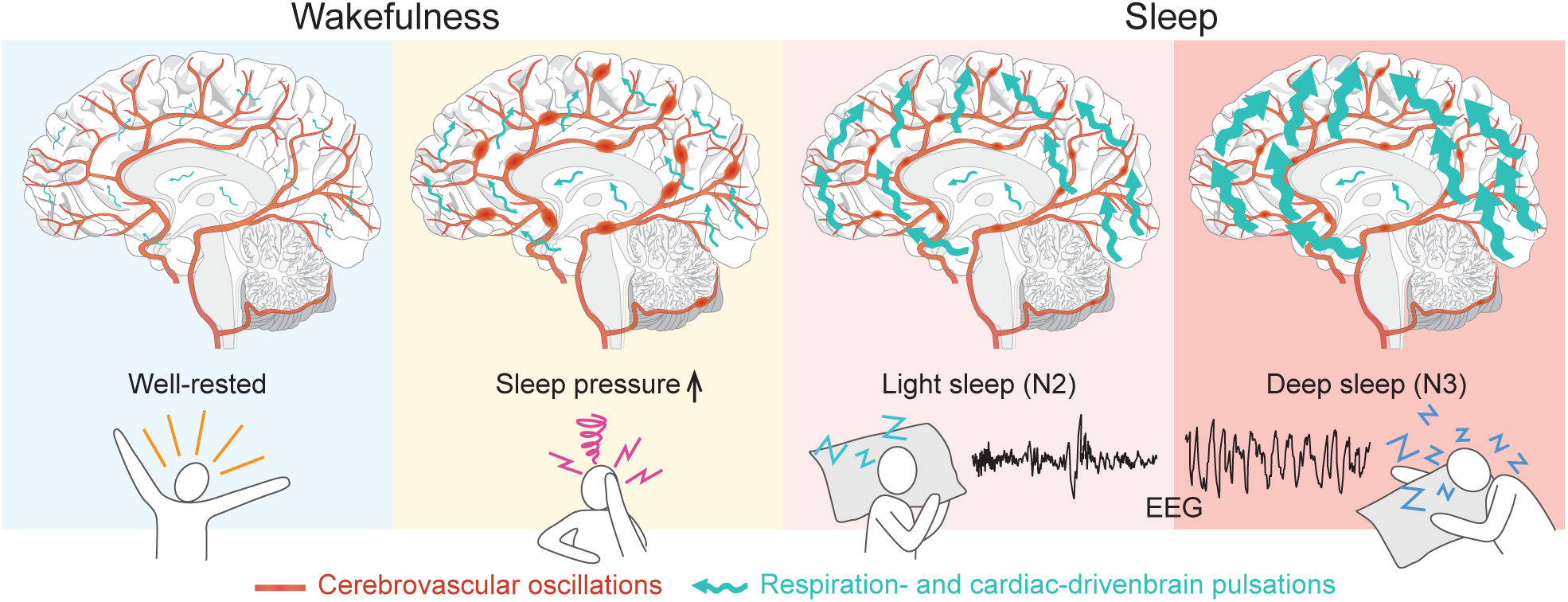

## Introduction

Deep sleep is essential for maintaining healthy brain function. A prominent characteristic of deep non-rapid eye movement (NREM) sleep is the presence of slow, homeostatically regulated, high amplitude waves in the electroencephalogram (EEG) delta range (0.5 to 4.5 Hz)^1^. These EEG slow waves are increased by sleep deprivation^2,3^ and have been implicated in multiple basic physiological processes, including brain plasticity^4^, memory consolidation^5^ and metabolic and immune system function^6^. Emerging evidence from animal studies indicates that EEG slow waves also play an important role in brain fluid dynamics^7^; the exchange of cerebrospinal fluid (CSF) and interstitial fluid (ISF) is regulated by NREM sleep, which promotes the influx of CSF from perivascular spaces into the brain parenchyma and causes a substantial increase in interstitial space volume^8–10^. Moreover, CSF-influx into the parenchyma strongly correlates with EEG delta power, which is a measure of slow wave activity^9,11^. This sleep-dependent exchange of CSF and ISF facilitates the clearance of metabolic waste from the rodent brain^8,12,13^, and has been termed ‘the glymphatic system’^14^.

Glymphatic flow is driven by an interplay of low frequency cerebrovascular oscillations, respiration and the cardiac cycle. Respiratory and cardiac rhythms generate pressure waves that physically propel CSF into the parenchyma^15,16^, while cerebrovascular oscillations, or vasomotion, not only propel CSF-flow along periarterial spaces^17,18^, but also help regulate it. This is evidenced by recent studies demonstrating that the sleep-wake cycle and neural activation affect both the constriction/dilation dynamics of the brain vasculature and the size of and flow within perivascular spaces^18–20^. The critical role of physiological drivers for glymphatic flow is underscored by pharmacological studies, demonstrating that enhancing arterial pulsatility via systemically administered adrenergic agonist directly affects both CSF-flow within perivascular spaces and CFS-ISF exchange throughout the mouse brain^21,22^.

Emerging evidence supports the facilitatory role of sleep in CSF-ISF exchange and brain clearance mechanisms in humans. A night of sleep is associated with increases in brain diffusivity and CSF volume^23,24^, while light NREM sleep enhances CSF inflow into the fourth ventricle^25^. Consistent with this, diffusivity in white matter regions of the brain decreases from morning to evening^26^, and sleep deprivation causes a reduced clearance of contrast agent from the brain^27^ as well as an increased accumulation of tau and β-amyloid in the CSF and brain parenchyma^28–30^. In further alignment with findings from rodent studies, intracranial pressure (ICP) measurements of the so-called B-waves (0.5 – 2 waves pr min), which are particularly prominent when ICP increases and are thought to reflect cerebrovascular oscillations^31^, are also influenced by sleep stage^32,33^.

The novel ultra-fast brain imaging technique Magnetic Resonance Encephalography (MREG)^34^ has emerged as a valuable tool for investigating brain-fluid dynamics in humans. MREG can achieve temporal resolutions on the order of milliseconds, allowing for detailed analysis of dynamic brain processes. That is, it enables non-invasive assessments of low frequency brain oscillations (LFOs) as well as respiration- and cardiac-driven brain pulsations^35,36^. Previous studies have shown that NREM sleep (pooled N1 and N2) enhances all three types of physiological brain oscillations^37^, with sleep-effects encompassing larger brain regions in N2 than N1 sleep^38^. Moreover, sleep deprivation seems to increase LFOs in N1 sleep^38^. Although the influence of circadian factors and sleep propensity on these findings remains unclear, they indicate that MREG measurements reflect brain fluid flow in humans.

Our study aimed to establish how physiological modulations influence MREG-detected brain oscillations, and how the strength of these brain oscillations correlates with sleep pressure, NREM sleep, sleep depth and EEG slow waves, while controlling for circadian rhythm, time-of-day and caffeine intake. First, we assessed the causal link between changes in MREG-detected brain oscillations and modulations of cerebrovascular oscillations, respiration, and cardiac activity by examining healthy individuals who, during MREG scans, performed either a breath-hold or the Valsalva manoeuvre, which is known to increase ICP^39,40^ and dampen cardiovascular pulsatility^41^. Next, we assessed the association between simultaneously acquired MREG and EEG signals in healthy individuals during well-rested wakefulness, after being kept awake for 35 hours and in NREM sleep stages N2 and N3. Moreover, we examined the effect of pharmacologically induced vasodilation through a randomized, double-blind, placebo-controlled, cross-over administration of the α1 and β-adrenergic antagonist carvedilol.

We hypothesized that 1) sleep deprivation is associated with a homeostatic drive towards enhanced physiological brain oscillations, 2) NREM sleep further enhances these oscillations, 3) the strength of physiological brain oscillations is proportional to sleep depth and EEG delta power, and 4) carvedilol modulates physiological brain oscillations.

## Results

### Effects of ICP, respiration and cardiac action on MREG spectral power

In four healthy individuals, we evaluated the effects of changes in ICP, respiration and cardiac activity on MREG-detected brain oscillations. Power peaks within physiological frequency ranges of the MREG spectrum were evaluated as measures of physiological brain oscillations. We observed at large increase in spectral power in a LFO frequency band (0.02 – 0.1 Hz) during the Valsalva manoeuvre, an established method to increase ICP^39,40^, compared to both normal breathing and breath-holding (Fig 1A&B). In 3 of 4 individuals, we also observed a slight increase in LFO spectral power during breath-holding, which increases brain blood volume^42,43^ and consequently, to some extent, ICP (Fig S1). Consistent with expectation, our data also confirmed that breath-holding and the Valsalva manoeuvre, which both pause respiration, cause a suppression of spectral power peaks in respiratory frequencies (0.14 - 0.5 Hz; Fig 1A&C). For the cardiac-related peaks in the MREG spectra (0.68-2.0 Hz), we observed a reduction in spectral power during the Valsalva manoeuvre, which is also known to lower stroke volume, blood pressure, and cerebral perfusion pressure^40,41^ (Fig 1A&D). Unless otherwise specified, these patterns were observed across all participants (Fig S1).

**Fig. 1.**
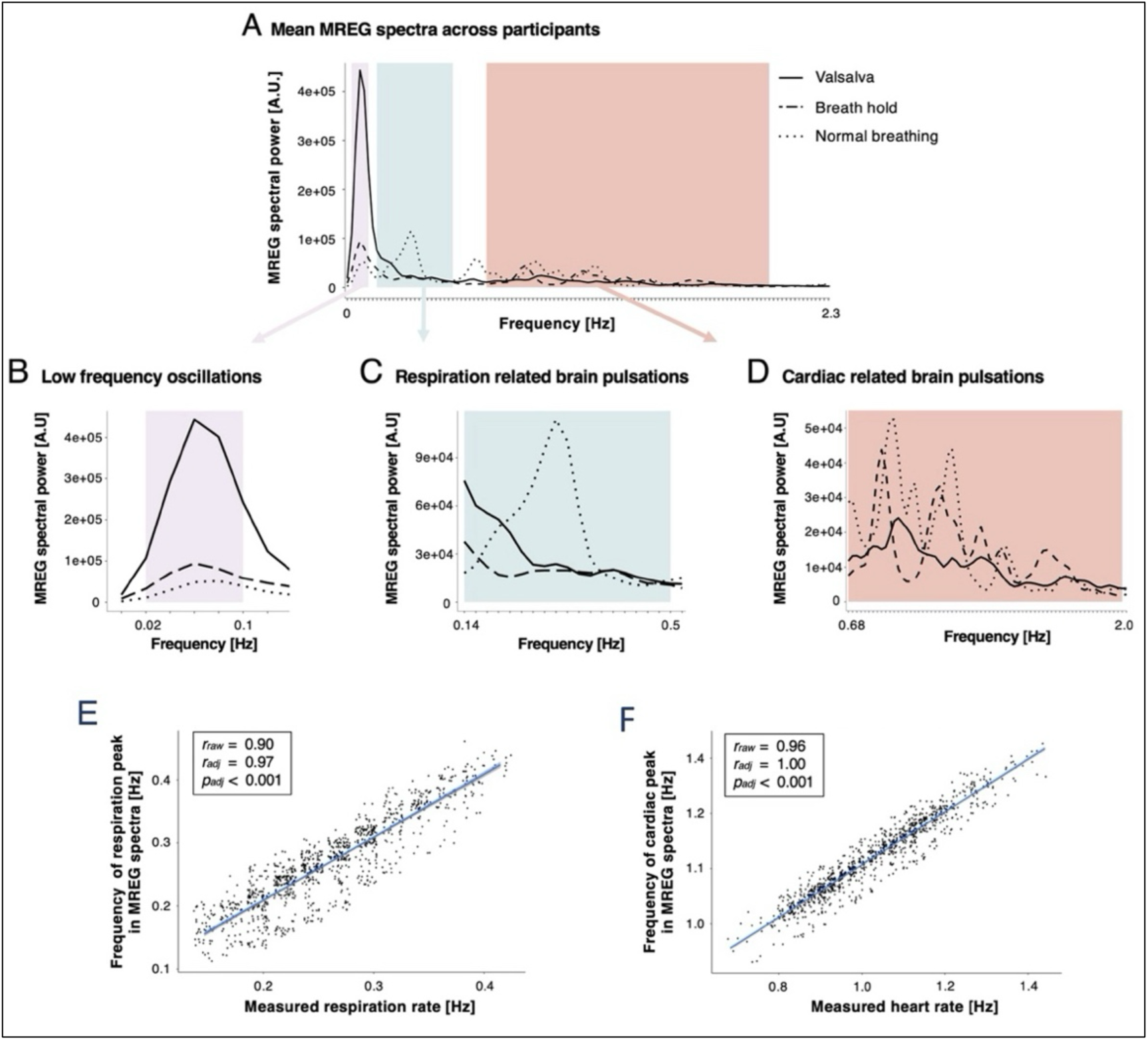
MREG-detected brain oscillations are affected by physiological modulation of intracranial pressure, respiration and cardiac action. (A) Mean whole-brain MREG spectra during normal breathing, breath-holding and the Valsalva manoeuvre, averaged across 3 sessions and 4 healthy individuals. Shaded areas represent the physiological frequency ranges in which power peaks were evaluated as measures of physiological brain oscillations. These were: (B) Low frequency oscillations (0.02 – 0.1 Hz), (C) respiration-related brain pulsations (0.14 – 0.50 Hz = 8.4 – 30 min^-1^), and (D) cardiac-related brain pulsations (0.68 – 2.0 Hz = 41 – 120 min^-1^). Lines represent group means across the three conditions. (E, F) Correlations between measured respiration rates and the frequency of respiration-associated peaks in the MREG spectra and between measured heart rates and the frequency of the cardiac-associated peak in the MREG spectra from all 30 sec epochs in the well-rested scans in the sleep study (*N* = 20).

Wakefulness data from the sleep trial (*N* = 20, described below) also showed a strong correlation between measured respiration rates (*r_raw_* = 0.90, *r_adj_* = 0.97, *p_adj_* < 0.001) and heart rates (*r_raw_* = 0.96, *r_adj_* = 1.00, *p_adj_* < 0.001), and the frequency of the corresponding peaks in the MREG spectra (Fig 1E&F).

### Sleep study cohort and descriptives

In a sleep trial, including a randomised double-blind, placebo-controlled, cross-over administration of the adrenergic antagonist carvedilol (Fig 2), we investigated the effects of sleep deprivation, deep NREM sleep and adrenergic inhibition on physiological brain oscillations in 20 healthy males with anamnestic and polysomnographically verified normal sleep (Table 1). Three MR/EEG scans sessions were conducted after an average of 11 (well-rested) and 35 hours (supervised sleep-deprived; placebo and carvedilol) of wakefulness (Table S1). All participants adhered to their sleep-wake schedules, as verified by actigraphy and sleep diary, and wakefulness during sleep deprivation periods was confirmed with continuous EEG recordings (Table S1). Sleep patterns across the three standardised nights leading up to scan days were comparable (Table S2). Participants were mostly awake during well-rested scans, but once sleep-deprived, they quickly entered NREM sleep stages N2 and N3 (Fig S2).

**Fig. 2.**
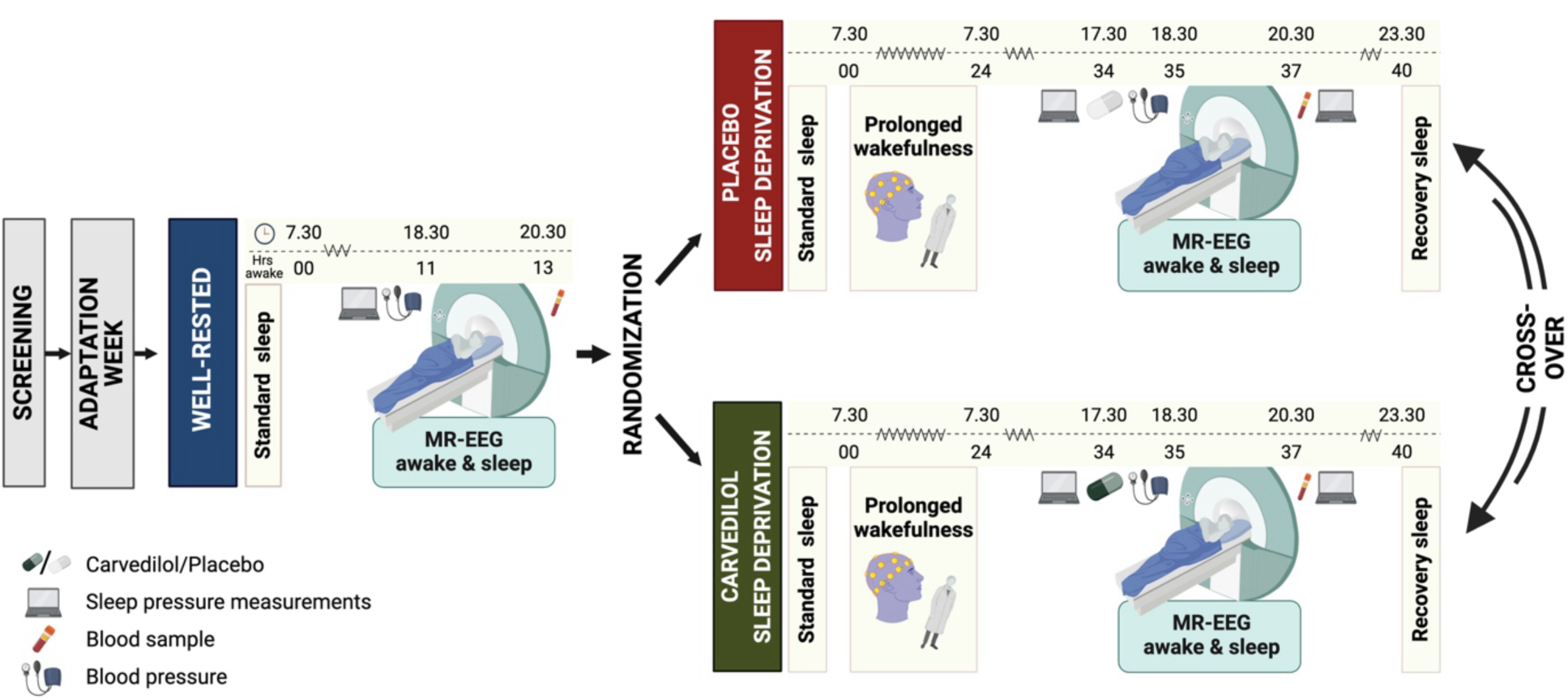
Study design of the sleep study. Setup and timings for the three study sessions. A well-rested and two sleep-deprived study sessions were conducted in the evenings at similar circadian time points. All three MR/EEG scans included a period of wakefulness followed by a sleep opportunity. Before the two sleep-deprived scans, participants received either a placebo or 25 mg of the adrenergic antagonist carvedilol in a double-blind, randomized, cross-over manner. *N* = 20. *Figure created with BioRender.com*.

**Table 1.** Demographics for sleep study cohort. BMI: Body mass index, MEQ: Morningness-eveningness questionnaire, ESS: Epworth Sleepiness Scale, PSQI: Pittsburgh Sleep Quality Index.

|  | Healthy volunteers<br>(N = 20) |
| --- | --- |
| **Age (years)** |  |
| Mean ± SD | 24.1 ± 2.8 |
| [Min, Max] | [20, 29] |
| **BMI (kg/m<sup>2</sup>)** |  |
| Mean ± SD | 22.7 ± 2.8 |
| [Min, Max] | [19.1, 31.6] |
| **Reported habitual sleep duration (h)** |  |
| Mean ± SD | 7.6 ± 0.4 |
| [Min, Max] | [7.0, 8.0] |
| **Morningness-eveningness (MEQ)** |  |
| Mean ± SD | 48.6 ± 6.4 |
| [Min, Max] | [38.9, 61.0] |
| **General daytime sleepiness (ESS)** |  |
| Mean ± SD | 6.2 ± 3.0 |
| [Min, Max] | [2.0, 13] |
| **Sleep quality index (PSQI)** |  |
| Mean ± SD | 3.0 ± 1.6 |
| [Min, Max] | [1.0, 6.0] |

Heightened sleep propensity due to sleep deprivation was confirmed by an increase in awake EEG delta power during the sleep-deprived scans (delta to total power ratio: 0.033 ± 0.017 AU, *p* = 0.051, *N* = 20) and impaired psychomotor vigilance task (PVT) performance before the scan sessions (reaction time: -0.035 ± 0.005 sec; lapses of attention +3.86 ± 0.23; *p*_both_ < 0.001, *N* = 20). As expected, NREM sleep was associated with a strong increase in EEG delta power (0.081 ± 0.014 AU, *p* < 0.001, *N* = 20) compared to sleep-deprived wakefulness, with N3 exhibiting higher EEG delta power than N2 sleep (*p* < 0.001, *N* = 17), and showed a small drop in respiration (-9%, *p* = 0.01) and heart rate (-11%, *p* < 0.001) (Table S3).

Treatment blinding was successful; participants guessed whether they received carvedilol or placebo before the two sleep-deprived scans only at chance level (guess accuracy: 54%).

### Sleep deprivation enhances spectral power in the LFO frequency band

Effects of sleep deprivation and slow-wave-rich NREM sleep (combined stages N2 & N3) on whole-brain LFOs were evaluated by assessing MREG spectral power in a frequency band similar to that of B-waves (0.012 – 0.034 Hz; ∼ 0.7 – 2 waves pr min). Analyses included 5-min scans from well-rested and sleep-deprived placebo conditions (Table S5; *see methods*).

Sleep deprivation caused a 120% increase in awake spectral power within the LFO frequency band (estimated mean in rested (10^6.748^) versus sleep-deprived (10^7.091^) wakefulness, 120% = (10^7.091^– 10^6.748^) /10^6.748^, *p* = 0.030; Fig 3B). During sleep-deprived wakefulness, spectral power of LFOs was also numerically higher than during subsequent periods of sleep-deprived NREM sleep (+ 49%, *p* = 0.19; Fig 3A-C).

**Fig. 3.**
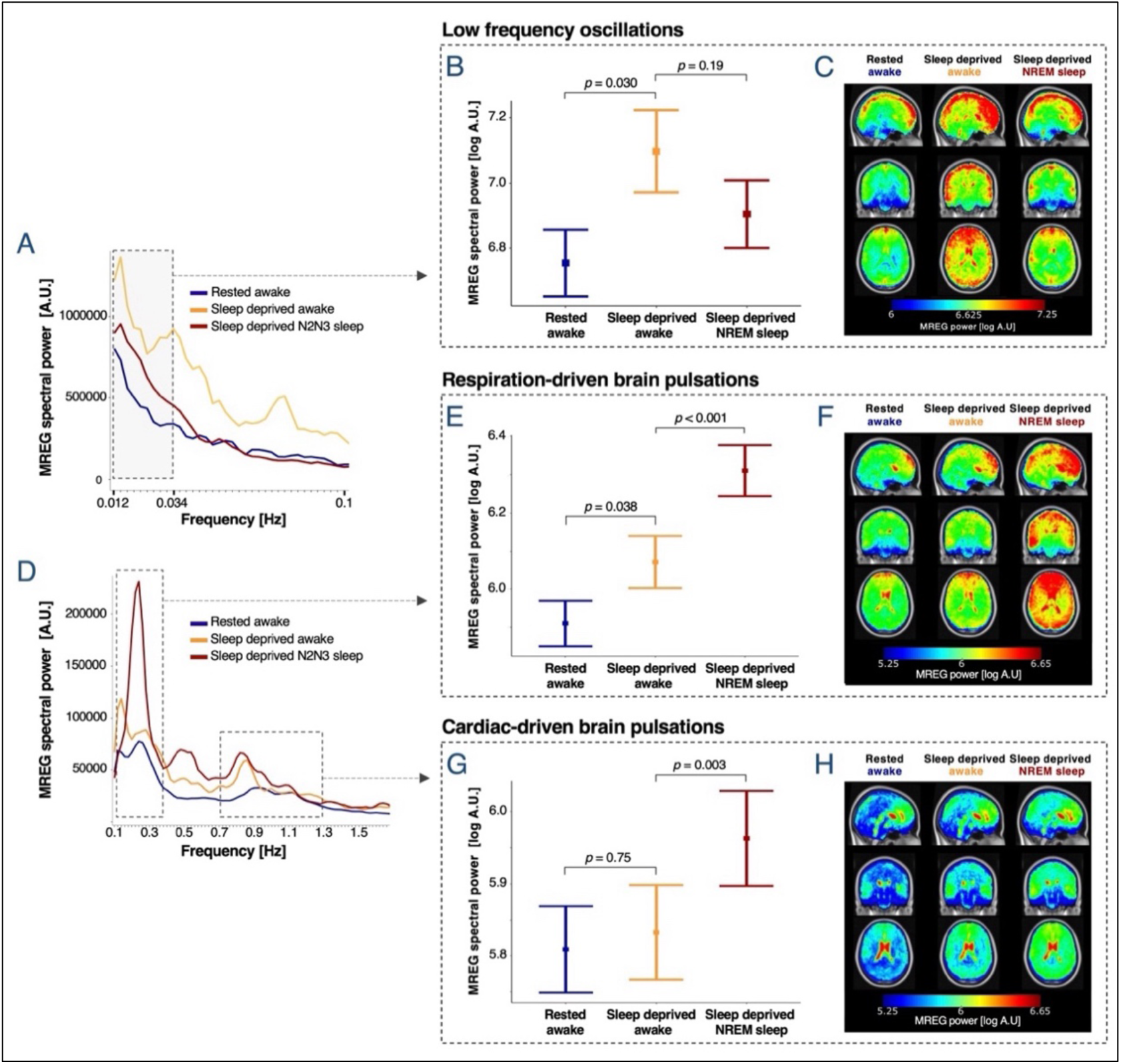
Sleep deprivation increases LFOs, while NREM sleep is the primary enhancer of respiration- and cardiac-driven brain pulsations. **(A**, **D)** Mean whole-brain MREG spectra across participants. Dashed boxes denote frequency ranges for LFOs (0.012-0.034 Hz) and for respiration- and cardiac-related peaks (individual frequency ranges were tailored epoch-wise from measured respiration and heart rates). **(B**, **E**, **G)** Estimated means ± SEM and *p*-values are from linear mixed models evaluating effects of sleep deprivation and sleep on spectral power in LFO (B), respiration (E), and cardiac (G) frequency bands. **(C**, **F**, **H)** Whole-brain power maps illustrates voxel-wise spectral power [log A.U.] in the LFO, respiration and cardiac frequency bands across all 5-min scans (LFO) or 30-sec epochs (respiration and cardiac), averaged across first epochs/scans, then participants (warped into MNI space). *N* included in analyses = 20 (see Tables S3 and S5 for number of participants in the three conditions).

To investigate the relationship between increased sleep pressure and LFOs, we then examined the association between LFO spectral power and psychomotor vigilance, as quantified with the PVT test before and immediately after scans sessions. We observed a correlation between LFO spectral power during sleep-deprived NREM sleep and pre-scan PVT performance (lapses of attention: *r_adj_ =* 0.50, *p =* 0.018; reaction time: *r_adj_* = 0.49, *p =* 0.023; Fig 4A-B). Improvement in PVT performance after having had a nap in the scanner (i.e., shorter reaction time and fewer lapses) also correlated positively with strength of LFO spectral power during NREM sleep (Δlapses: *r_adj_* =-0.48, *p =* 0.015; Δreaction time: *r_adj_* = -0.39, *p =* 0.043; Fig4C-D). We observed no evidence for such correlation during wakefulness (*p_all_* > 0.5).

**Fig. 4.**
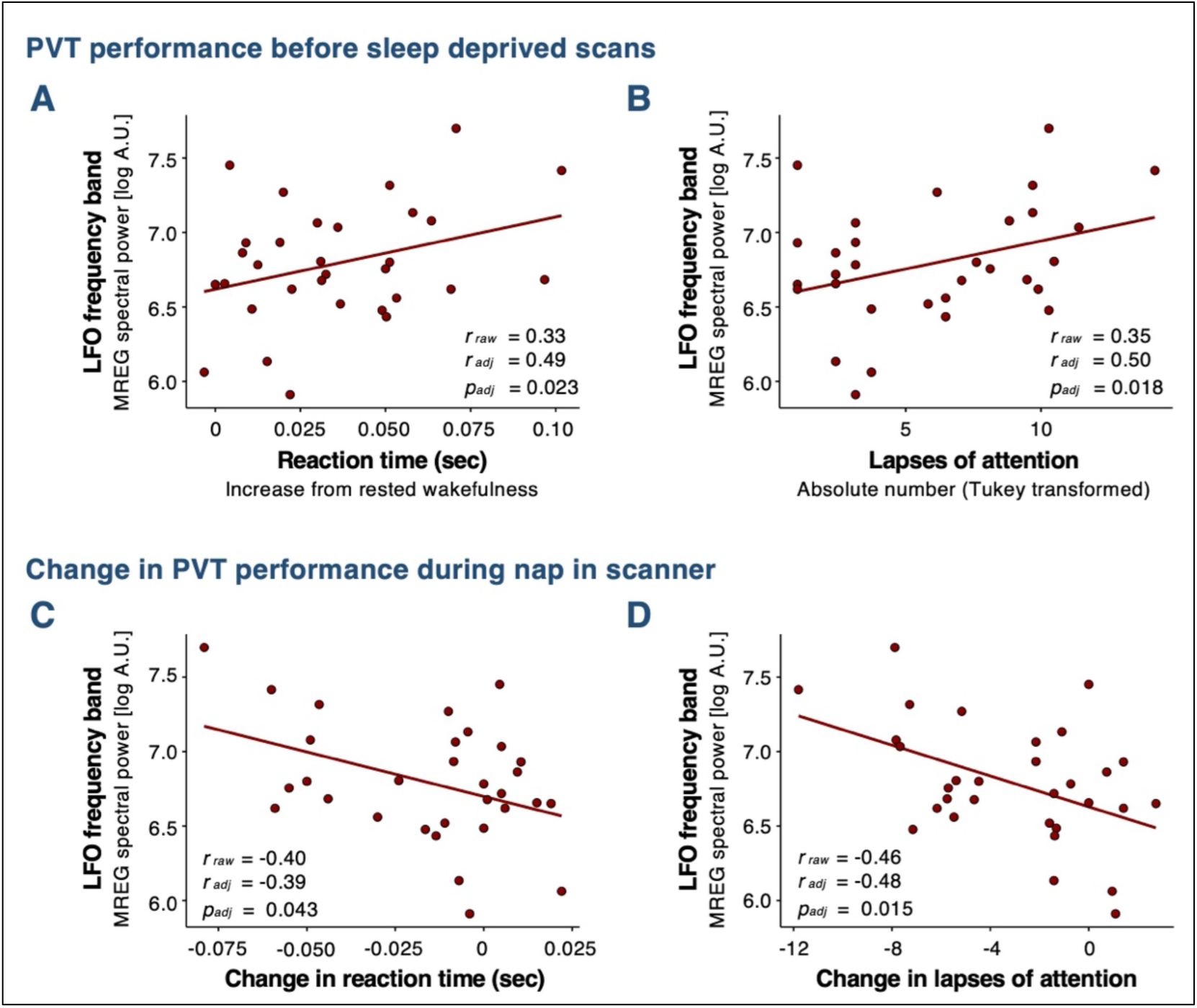
PVT performance is associated with LFOs during sleep. **(A**, **B)** Correlations between pre-scan measures of impaired PVT performance (i.e. longer reaction times and more lapses of attention) and intensity of LFO spectral power during sleep. **(C, D)** Improvement in PVT performance (i.e. lowering of reaction time and less lapses) from before to after nap in scanner correlates with strength of LFO spectral power during sleep. Data from sleep deprivation scans, from both placebo and carvedilol sessions, are presented. Mean LFO spectral power for scan was calculated for each participant (for both scans) from all 5-min periods classified as NREM sleep (stages N2 or N3). *r*_raw_ denotes Pearson’s corelation coefficient. *r*_adj_ and *p*_adj_ denote adjusted estimates from linear mixed models. *N*_placebo_ = 14, *N*_carvedilol_ = 17.

Given earlier findings of increased LFOs during fully or partly sleep-deprived N1-N2 sleep^25,37,38^, we next assessed the difference between rested wakefulness and sleep-deprived N2-N3 sleep, without controlling for the embedded effects of sleep deprivation. Here, we observed a non-significant 48% increase in LFOs by sleep (*p* = 0.21, Fig 3B).

### NREM sleep enhances MREG spectral power in the respiration and cardiac frequency bands; N3 more so than N2 sleep

To investigate how sleep deprivation, vigilance state and sleep depth affect respiration- and cardiac-driven brain pulsations, we assessed MREG spectral power within individually tailored respiration- and cardiac frequency bands (see methods) in 30-sec epochs from rested and sleep-deprived placebo scans (Table S3 & S6).

Compared to rested wakefulness, awake sleep-deprived participants had increased whole-brain spectral power within the respiration (+45%, *p* = 0.038, Fig 3D, E-F), but not in the cardiac frequency band (+6%, *p* = 0.75, Fig 3D, G-H). When participants fell asleep and entered NREM sleep (N2 and N3 sleep combined), we observed a strong increase in whole-brain spectral power in both the respiration (+73%, *p* < 0.001, Fig 3D, E-F) and cardiac frequency bands (+35%, *p* = 0.003, Fig 3D, G-H). The sleep-induced increase in spectral power was highly consistent across participants and were identified in 15 out of 16 participants in the respiration frequency band and in 12 out of 17 participants in the cardiac frequency band.

Sleep depth also modulated MREG spectral power. In the respiration frequency band, N2 sleep was associated with a 35% increase in spectral power compared to sleep-deprived wakefulness (*p*_adj_ = 0.047), while N3 sleep further increased spectral power by 57% (*p_adj_* < 0.001) (Fig 5A-C). In the cardiac frequency band, spectral power was higher during N3 sleep than during both N2 sleep (+39%, *p_adj_* < 0.001) and sleep-deprived wakefulness (+58%, *p_adj_* < 0.001). A numerical, but not statistically significant, increase in spectral power from sleep-deprived wakefulness to N2 sleep was also observed (+13%, *p_adj_* = 0.19) (Fig 5A & D-E).

**Fig. 5.**
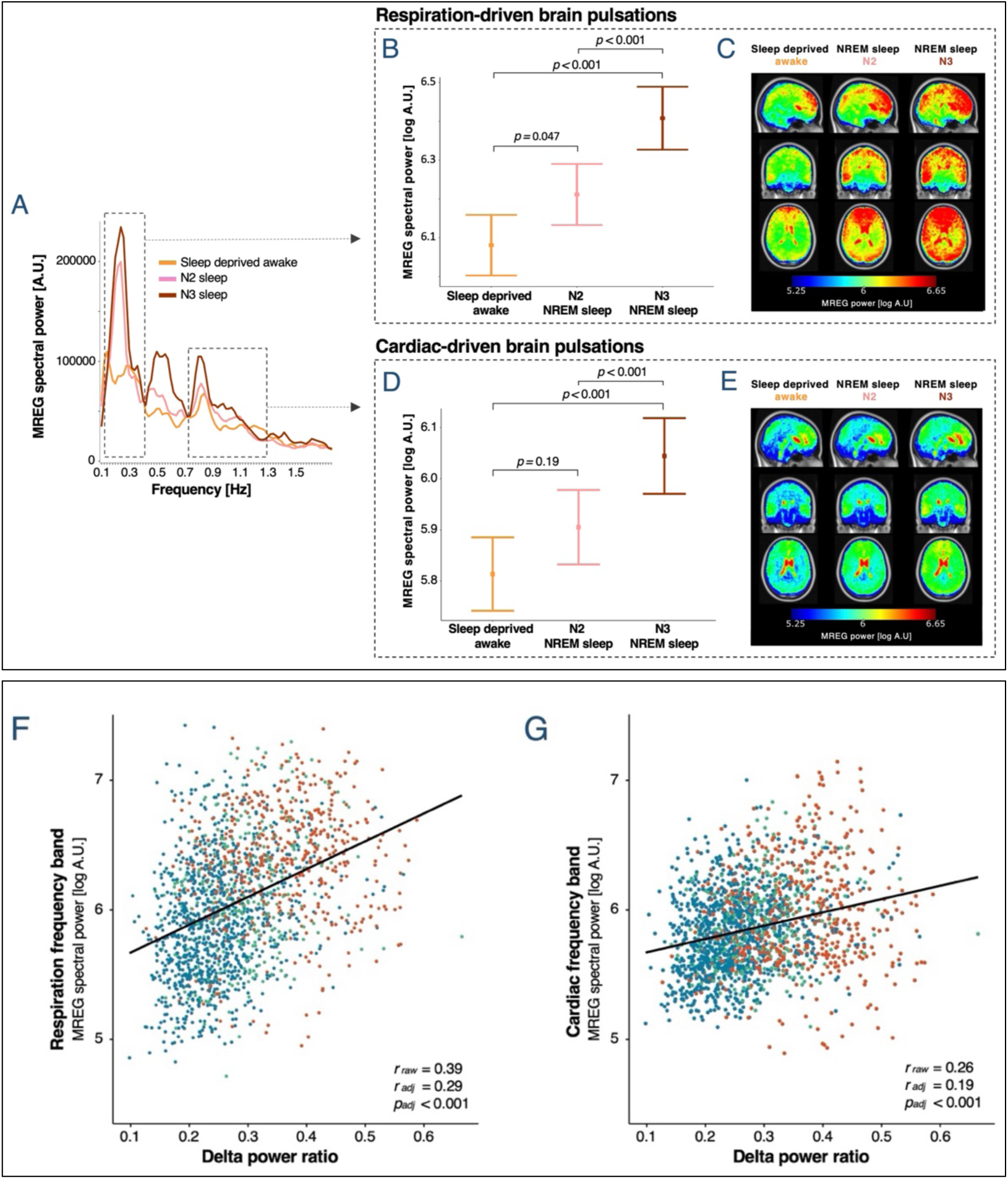
Strength of respiration- and cardiac-driven brain pulsations correlates with sleep intensity. **Upper panel: (A)** Mean whole-brain MREG spectra across participants for sleep-deprived wakefulness, N2 sleep and N3 sleep (placebo scans). Dashed boxes denote frequency ranges, in which respiration- and cardiac-related peaks are found (individual frequency ranges were tailored epoch-wise from measured respiration and heart rates). **(B**, **D)** Estimated mean ± SEM and p-values are from linear mixed models evaluating the effect of sleep stage on spectral power in respiration and cardiac frequency bands. All p-values have been adjusted for multiple comparison with Bonferroni correction. **(C, E)** Whole-brain power maps illustrate voxel-wise spectral power [log A.U.] in the respiration and cardiac frequency bands across all 30-sec epochs, averaged across first epochs, then participants (warped into MNI space). *N* = 19 (see Table S6 for number of participants in each of the conditions). **Lower panel:** EEG delta power ratio versus simultaneously acquired spectral power in the respiration **(F)** and cardiac **(G)** frequency bands in all 30-sec epochs from both rested and sleep-deprived placebo scans. Colours represent the scored sleep stages (blue: rested wakefulness; red: NREM sleep stage 2 and 3; green: uncertain scorings) and are included for visualisation, but were not included in analysis*. r*_raw_ denotes Pearson’s corelation coefficient. *r*_adj_ and *p*_adj_ denote adjusted estimates from linear mixed models. *N_resp_* = 19 and *N_card_* = 20.

We did not observe any correlation between PVT performance and spectral power in the respiration or cardiac frequency bands.

### EEG delta-power during MR-imaging correlates with MREG spectral power in respiration and cardiac frequency bands

To validate the found effects of slow-wave-rich NREM sleep and sleep depth on respiration- and cardiac-driven brain pulsations in a quantitative and sleep-stage independent manner, we quantified EEG delta-power ratio across all 30-sec epochs from well-rested and sleep-deprived placebo scans, irrespective of their scored vigilance state (Fig 5, lower panel). This showed a positive correlation between EEG delta power ratio and whole-brain spectral power in both the respiration (*r_raw_* = 0.39, *r_adj_* = 0.27, *p_adj_* < 0.001; Fig 5F) and cardiac (*r_raw_* = 0.26, *r_adj_* = 0.19, *p_adj_* < 0.001; Fig FG) frequency bands. This positive association was identifiable in most participants (Fig S3).

### NREM sleep primarily enhances MREG spectral power in the respiration and cardiac frequency bands in grey and white matter

To explore whether the strength of physiological brain oscillations differed between grey matter (GM), white matter (WM) and CSF, we analysed data from well-rested and sleep-deprived placebo scans. During rested wakefulness, LFO spectral power was found to be similar in GM, WM and CSF (Fig 6A). This similarity persisted after sleep deprivation, where LFO spectral power increased to a similar extent in all tissue types (‘tissue type’ × ‘sleep deprivation’ interaction: *p =* 0.96; Fig S4A). Additionally, we did not observe any interactions between NREM sleep and tissue type for LFO spectral power (*p =* 0.92; Fig 6B&C).

**Fig. 6.**
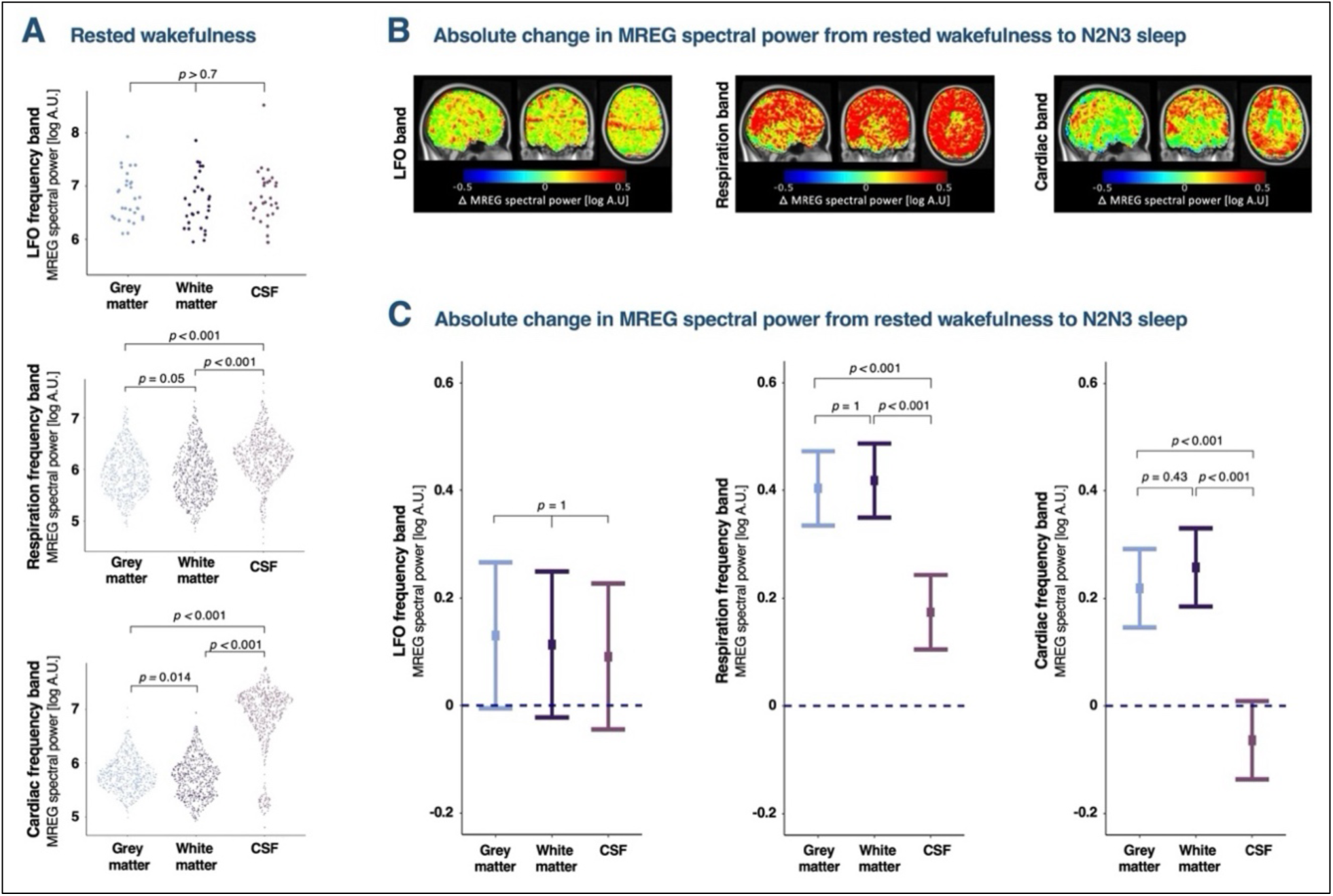
Sleep has a greater effect on respiration- and cardiac-driven brain pulsations in grey and white matter than in CSF. **(A)** Difference in brain oscillations strength between grey matter, white matter and CSF in ventricles in rested wakefulness. Each data point represents a 30-sec epoch from a single participant. **(B)** Voxel-wise power maps illustrate the averages of participant-wise absolute differences in spectral power between rested wakefulness and NREM sleep (stages N2 and N3) in LFO, respiration on cardiac frequency bands. Whole-brain spectral power maps of individually calculated voxel-wise differences were warped into MNI space and averaged across participants. **(C)** Changes in estimated means of MREG spectral power (log10) in LFO, respiratory and cardiac frequency bands from rested wakefulness to N2/N3 sleep. *p*-values represent interaction effects between tissue type and sleep from linear mixed models and are adjusted for multiple comparisons with Bonferroni correction. *N* = 20 (see Tables S3 and S5 for number of participants in each condition).

By contrast, in the respiration and cardiac frequency bands, spectral power varied significantly across CSF, GM and WM (‘tissue type’ main effect: *p* < 0.001). During rested wakefulness, CSF showed notably higher spectral power than both GM (respiration band: +101%, *p_adj_* < 0.001; cardiac band: +1030%, *p_adj_* < 0.001) and WM (respiration band: +118% *p_adj_* < 0.001; cardiac band: +1153%, *p_adj,_* < 0.001). Additionally, participants also had higher spectral power in GM than in WM in the well-rested condition (respiration band: +9%, *p* = 0.017, *p_adj_* = 0.052; cardiac band: +11%, *p* = 0.005, *p_adj_* = 0.014) (Fig 6A).

The effect of NREM sleep on respiration- and cardiac-driven brain pulsations was more pronounced in GM and WM than in CSF, with a significant interaction effect between vigilance state and tissue type (*p* < 0.001; Fig 6B&C). Specifically, GM and WM exhibited 70% and 75% stronger increases in spectral power than CSF in the respiration band, and 92% and 110% stronger increases than CSF in the cardiac band (*p_adj, all_* < 0.001).

All tissue types showed significant effects of NREM sleep on spectral power in both respiration- and cardiac frequency bands (respiration band_(all):_ *p* < 0.001; cardiac band_(all)_: *p* < 0.01). Additionally, sleep deprivation interacted with tissue type in the respiration band (*p* < 0.001), with effects confined to GM (*p* = 0.045) and WM (*p* = 0.054) (Fig S4 B&C).

### Adrenergic antagonism decreases spectral power in LFO and cardiac frequency bands

Treatment with the adrenergic antagonist carvedilol demonstrated anticipated systemic effects: it reduced serum norepinephrine levels ^44^ (-512.5 ± 129.9 pg/mL, *p_adj_* = 0.002, *N* = 19; Fig S5), lowered mean arterial blood pressure by ∼ 5 % (-4.4 ± 0.9 mmHg, *p* < 0.001, *N* = 20), and had no significant effect on either heart or respiration rates (Table S3). Carvedilol did not change absolute or relative time spent in wakefulness or sleep during scans (Fig S2) and only had minor effects on recovery sleep (Table S4).

During NREM sleep, carvedilol was associated with a decrease in LFO spectral power (-51%, *p* = 0.050), while its effect during wakefulness was only numerically negative (-35%, *p* = 0.32) (Fig 7A-C). Similarly, carvedilol significantly reduced spectral power in the cardiac frequency band during NREM sleep (-30%, *p* = 0.032), but not wakefulness (Fig 7D, G-H). In the respiration frequency band, carvedilol intervention showed comparable spectral power outcome as placebo during both sleep-deprived wakefulness and NREM sleep (‘treatment’: *p* = 0.56, ‘NREMsleep_carvedilol_’: *p <* 0.001, Fig 7D&E-F).

**Fig. 7.**
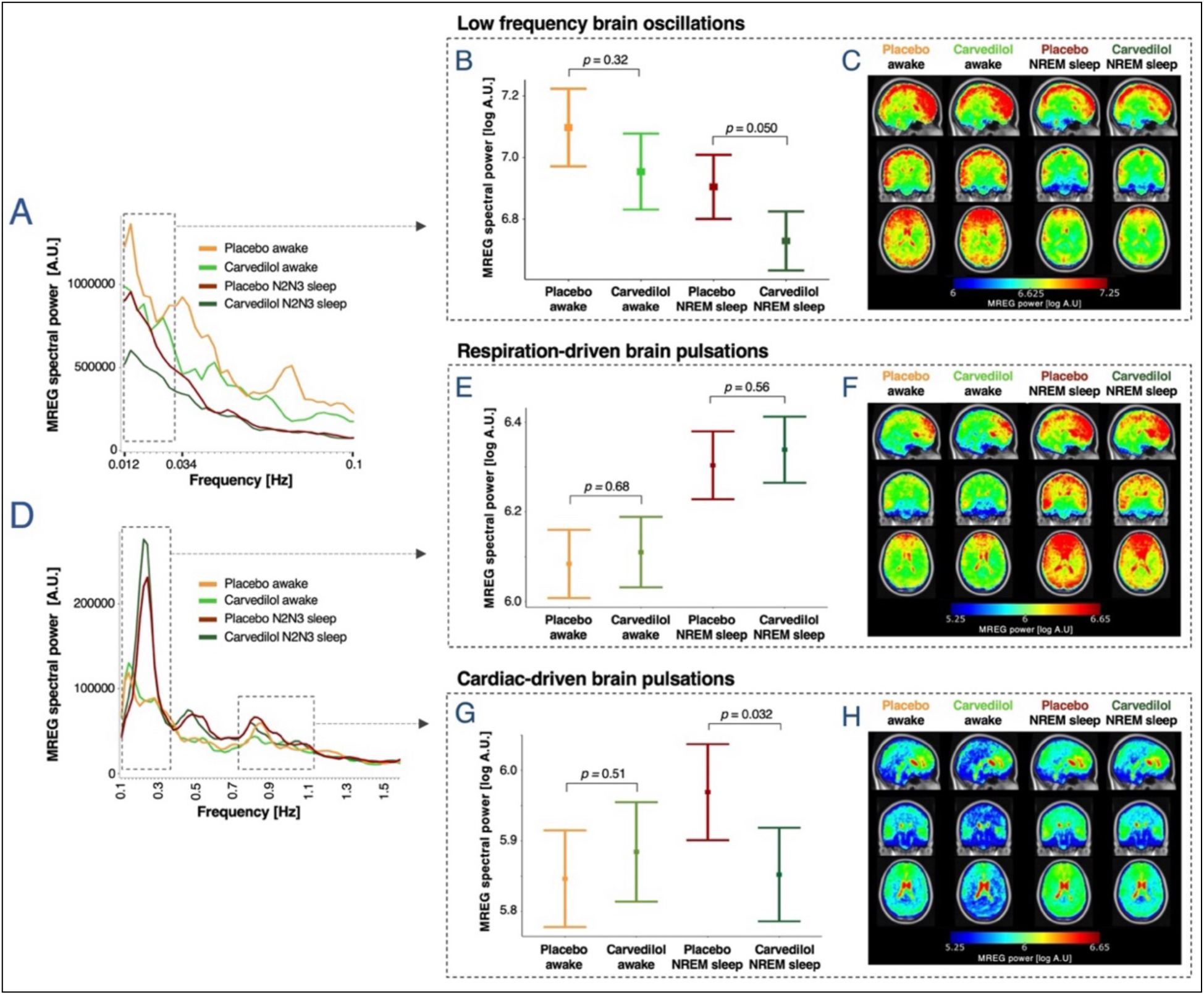
The α1 and β-adrenergic antagonist carvedilol attenuates LFOs and cardiac-driven brain pulsations during sleep. **(A, D)** Mean whole-brain MREG spectra across participants in the sleep-deprived scans for placebo wakefulness, placebo NREM sleep, carvedilol wakefulness and carvedilol NREM sleep. Dashed boxes denote frequency ranges for LFOs (0.012-0034 Hz) and for respiration- and cardiac-related peaks (individual frequency ranges were tailored epoch-wise from measured respiration and heart rates). **(B, E, G)** Estimated means +/- SEM and *p*-values are from linear mixed models evaluating the effect of treatment and NREM sleep on spectral power in the LFO, respiration and cardiac frequency bands. **(C**, **F**, **H)** Whole-brain power maps illustrate voxel-wise MREG spectral power [log A.U.] in the LFO, respiration and cardiac frequency bands across participants, averaged across first all 5-min scans (LFO) or 30-sec epochs (respiration and cardiac) per participant, then across participants (warped into MNI space). *N* = 20 (see Tables S3 and S5 for number of participants in each of the three conditions).

## Discussion

In this circadian-controlled sleep and sleep deprivation study, we demonstrate that heightened sleep pressure promotes low frequency brain oscillations, while NREM sleep depth and EEG delta power correlate with the strength of respiration- and cardiac-driven brain pulsations. Moreover, we show that LFOs are affected by the modulation of vascular pulsatility through adrenergic antagonism, supporting that these originate from cerebrovascular oscillations, or vasomotion.

First, we establish a causal link between changes in MREG-detected brain oscillations and modulations of cerebrovascular oscillations, respiration and cardiac activity. We show that a dynamic increase in ICP from the Valsalva manoeuvre^39,40^ leads to a prominent increase in MREG spectral power in frequencies below 0.1 Hz. As this effect mirrors the behaviour of ICP monitored B-waves (0.01 – 0.33 Hz), which increase with rising ICP and are thought to reflect cerebrovascular oscillations^31^, we propose that MREG-detected LFOs reflect cerebral vasomotion. Additionally, we validate that MREG-detected brain pulsations in cardiac and respiratory frequency ranges are, in fact, causally linked to their corresponding physiological processes. This is evidenced by the elimination of respiration-related peaks in the MREG-spectra during breath-holding and the suppression of cardiac-related peaks during the Valsalva manoeuvre, which is known to reduce cardiac stroke volume and blood pressure. In further support of this causal relationship, we show that measured respiration and heart rates are closely correlated with the frequencies of peaks in the MREG-spectra in their respective frequency ranges.

The physiological interpretation of MREG-detected brain oscillations has previously been attributed either to the direct imaging of brain fluid motion, signifying CSF exchanges in humans^35,36^, or to imaged tissue oscillations reflecting the underlying brain tissue compliance, encompassing factors such as water content or tissue viscosity^45,46^. We suggest that these two mechanisms are intimately associated, as physiologically induced brain oscillations conceivably elicit changes in brain tissue water content as they drive CSF into the brain. Our data support this interpretation of the MREG signal for respiration- and cardiac-driven brain pulsations by demonstrating that, irrespective of vigilance state, these pulsations are significantly stronger in CSF than in grey and white matter. During rested wakefulness, they are also more pronounced in grey matter than in white matter. Considering that water content and viscosity vary across these three brain tissue types^47,48^, our results indicate that water content is directly associated with the strength of respiration- and cardiac-driven brain pulsations, and that increases in CSF influx to the brain parenchyma will lead to increases in these pulsations. For LFOs, we observed no differences in oscillation strength across tissues types, supporting the idea that these reflect global cerebral vasomotion and its associated effects on CSF inflow^7,25,49^, rather than parenchymal fluid flow or tissue compliance.

A novel observation of ours is that sleep deprivation has a greater effect on LFOs than sleep does. Specifically, MREG-detected LFOs increase on a whole-brain level during sleep-deprived wakefulness, compared to both rested wakefulness and subsequent periods of NREM sleep (stage N2 and N3). Moreover, we find that the strength of LFOs during sleep correlates with pre-scan measures of impaired vigilance attention (long reaction time and more lapses of attention), which is a well-validated indicator of homeostatic sleep drive^50,51^. When participants have a nap in the scanner, we also see a positive correlation between LFO strength and the improvement in psychomotor vigilance from before to after the nap. Collectively, these results suggest that sleep pressure is a strong driver of LFOs – and therefore cerebrovascular oscillations - and indicate that these oscillations play an important role for restoration of performance during sleep. This notion is supported by a recent MREG study, in which healthy individuals exhibited stronger LFOs during sleep-deprived N1 sleep than during rested N1 sleep^38^. While previous studies in humans have demonstrated that NREM sleep stages N1 and N2 increase the strength of LFOs^25,38,52^, they did not differentiate the effects of sleep deprivation from those of sleep itself, nor did they consider circadian influences or sleep pressure at scan onset. In the present study, we address these gaps. What we find is that LFOs are increased already in sleep-deprived wakefulness, whereby earlier reported influences of sleep on LFOs might potentially be obscured by sleep pressure effects. Sleep stage might also influence the magnitude of LFOs. ICP B-waves have been reported to occur more frequently in N1 and N2 sleep^32,33^ and less so during deeper sleep^53^, and similarly, both the slow oscillating inflow of CSF through the fourth ventricle and the simultaneously measured global signal have been reported to be strongest in N1 sleep, followed by N2 sleep, and weakest in N3 sleep^49^. Given that sleep pressure dissipates over time spent asleep^3^, this indicates that cerebrovascular oscillations are strongest at the beginning of sleep – when sleep pressure is at its peak and individuals typically reside in the lighter sleep stages. Of note, there were no significant changes in blood pressure, heart and respiration rates between well-rested and sleep-deprived wakefulness. Therefore, we do not believe that autonomic arousal alone can explain the observed increases in LFOs, as suggested by Picchioni and colleagues^49^. Taken together, ours and prior findings indicate that sleep pressure drives cerebrovascular oscillations, which are thought to be a key driver of CSF-ISF exchange and brain clearance.

Expanding on previous findings^38,52^, we show that deep NREM sleep (pooled N2 and N3 sleep) enhances respiration- and cardiac-driven brain pulsations at a global brain level, and as a novel finding, we show that the effect of sleep on these brain pulsations is more pronounced in grey and white matter than in CSF. As the strength of these physiological brain pulsations relates to tissue water content, we propose that this sleep-induced increase in pulsatility within the brain parenchyma supports a sleep-dependent increase in influx of CSF to the brain parenchyma and expansion of extracellular space volume in humans, similar to observations made in the mouse brain^8,10^.

Our study further demonstrates that the strength of both respiration- and cardiac-driven brain pulsations increases with the depth of NREM sleep, showing greater pulsatility in N3 than in N2 sleep. In line with this, we also show that the strength of these pulsations is positively correlated with EEG delta power, which is a measure of slow wave activity. A recent study of shallower NREM sleep^38^ agrees with these findings, reporting that respiration-driven pulsations increase only in N2 sleep (and not N1 sleep), while cardiac-driven pulsations encompass larger brain regions in N2 than in N1 sleep. The association between NREM slow waves – i.e. deeper sleep and higher delta power - and strength of brain pulsations may be explained by the synchronous neuronal activity of slow wave sleep driving fluid and solute movement physio-mechanically through the brain^11,54^. EEG delta power is also an established marker of homeostatic sleep propensity^2,55^, and its close association with respiration- and cardiac-driven brain pulsations, and thus brain-fluid flow, suggest that these may be homeostatically regulated and an integral function of human sleep. Our results closely mirror previous preclinical work demonstrating that CSF influx and perfusion through the brain parenchyma is associated with EEG slow waves^9,11^. Consequently, we here provide support for the hypothesis that humans - alike mice – have a sleep- and slow-wave dependent exchange of CSF and ISF.

Interestingly, we also noted an elevation of respiration-driven brain pulsations during sleep-deprived wakefulness, which was specific to grey and white matter. This observation suggests that when deprived of sleep, the brain actively engage in mechanisms to increase CSF influx to the brain parenchyma, albeit to a lesser extent than during NREM sleep. We speculate that this could represent an adaptive response to partially counteract accumulation of waste products.

The adrenergic antagonist carvedilol decreases heart contractility, lowers blood pressure, and inhibits smooth muscle contraction^56,57^, which putatively decreases vasomotor activity and cerebrovascular oscillations. Compared to placebo, this pharmacological modulation is associated with lower LFOs and cardiac-driven brain pulsations in NREM sleep. This finding strengthens the hypothesis that LFOs are generated by cerebrovascular oscillations and further underscores the causality between cardiac action and brain pulsations in the cardiac frequency band. As for the respiration-driven brain pulsations and carvedilol, we largely replicated observations made with placebo. Previous studies suggest that central inhibition of noradrenergic receptors increases CSF-ISF exchange^8,58^ and brain clearance^59^. However, our participants only received a single peroral dose of carvedilol, and given that such a dose of the treatment has been shown to enter the brain in only small amounts, possibly due to active ATP-dependent efflux mechanisms^57,60^, we believe that the major pharmacological effects took place on the vasculature. Consistent with this, we did not find any discernible differences in sleep patterns between placebo and carvedilol sessions. The notion is also consistent with preclinical work showing that systemic administration of adrenergic agonist increases the pulsatility of cortical arteries, which in turn drives CSF-influx to the brain^21,22^.

Our study is not without limitations. Firstly, to eliminate the influence of age and menstrual cycle on sleep and circadian rhythm^61^, we only included young males in the sleep study. While preclinical studies have not found any male/female differences in glymphatic flow and brain fluid dynamics, this choice limits the generalizability of our findings^62^. Secondly, there is no consensus in the literature clearly defining the appropriate frequency range for brain LFOs. We chose a range consistent with that previously defined for B-waves, but others have applied a broader frequency band when analysing human brain pulsations (∼ 0.01 - 0.1 Hz)^25,37^. In a posthoc analysis, we assessed if a LFO frequency band of 0.01 - 0.1 Hz would change our reported results and confirmed that the outcome was consistent with the 0.01 – 0.034 Hz band applied in our study. Finally, since we did not measure ICP directly, we cannot assess the correlation between ICP and the strength of LFOs directly. Regardless, this is the first study to carefully control for effects of sleep pressure, time of day, circadian rhythm, and alcohol and caffeine intake when evaluating the effects of vigilance state on MREG-detected brain pulsations.

### Conclusion

Our investigation unveils that, following 35 hours of wakefulness, LFO intensify throughout the entire brain and that subjective measures of sleep pressure exhibit a positive correlation with the magnitude of LFO during sleep. Additionally, our study reveals that adrenergic antagonism, which modulates vascular oscillations, dampens sleep-associated effects on LFO and cardiac-driven pulsations, providing compelling evidence supporting the notion that LFO represents vasomotor activity. These important findings offer crucial insights into how the vasomotor system in the brain responds to prolonged sleep deprivation, potentially serving as a means to mitigate the accumulation of brain waste products.

Furthermore, we illustrate that as sleep depth increases, brain pulsations induced by respiration and cardiac activity intensify in both grey and white matter. This observation aligns with preclinical evidence suggesting a sleep-dependent influx of CSF into the brain parenchyma. The strength of respiration- and cardiac-driven brain pulsations correlates with the depth of NREM sleep and EEG delta power, highlighting that EEG slow waves serve as indicators of the influx of CSF into the human brain. These new findings collectively contribute to our understanding of the intricate dynamics between sleep pressure and patterns, vasomotor activity, respiratory and cardiac actions on the brain, and the clearance of waste products from the brain.

## Materials and methods

### Study design for sleep study

We conducted a circadian-controlled sleep and sleep deprivation study that included a double-blind, placebo-controlled, cross-over administration of the adrenergic antagonist Carvedilol (Fig. 2). The study was preregistered at ClinicalTrials.gov (NCT03576664) and conducted at the Copenhagen University Hospital, Rigshospitalet, between January 2018 and May 2019. During an adaptation week and throughout the study period, participants followed a strict 8 h sleep, 16 h wake protocol. Sleep-wake rhythm was monitored continuously with actigraphy (Actiwatch spectrum, Philips Respironics) and a sleep diary to ensure that all measurements and assessments were performed during the same circadian phase and at comparable sleep propensities. Furthermore, participants were not allowed to drink alcohol and daily caffeine consumption was limited to two or less cups of coffee (≲ 200 mg caffeine/day) taken no later than 2 pm. As shown in Fig. 2, three EEG/MRI scan sessions were conducted on separate days in the evening (standardised time: ∼18.30-20.30): A well-rested scan scheduled after 11 hours of wakefulness followed by two sleep-deprived scans performed after 35 hours of wakefulness. Sleep-deprived scans were performed one week apart, with participants receiving either a placebo or 25 mg of the nonselective β1-, β2- and α1-adrenergic antagonist carvedilol one hour before scan start. Carvedilol reaches peak plasma concentration in 1-2 hours and has an elimination half-life of 4-7 hours^56^. Placebo and carvedilol were administered in identical capsules (manufactured and distributed by Capital Region Pharmacy), and participants were randomised to either the placebo-carvedilol or carvedilol-placebo sequence by a research administrator not otherwise involved in data collection or analysis. All three EEG/MRI scans sessions included a period of wakefulness (well-rested scan: ∼ 45 min; sleep-deprived scans: ∼30 min), followed by a one-hour sleep opportunity. During the awake period of the scans, lights were on, and we continuously checked that participants were awake. After the awake session, lights were turned off, the scanner environment was kept as silent as possible, and participants were instructed to relax and told that they were allowed to sleep. During this sleep opportunity, scans cycled between 5-min MREG- and multiband sequences to maintain a steady noise level within the scanner. After the lights-off session, lights were turned back on and participants were waken up. Participants’ blood pressure was measured before and immediately after MR scan sessions and a blood sample to quantify NE levels was collected from the participants ∼15 min after scans had ended. Sleep pressure was assessed with PVT performance before and after the scan sessions. During the nights leading up to each of the three study periods and in the two nights following the sleep deprivation periods, participants slept in a controlled sleep environment monitored with PSG (see supplements). The prolonged wakefulness periods were monitored with EEG and participants were continuously supervised by members of the research team to ensure that participants stayed awake throughout the wakefulness period. The sample size was based on data from Xie et al.^8^, which suggest a within-subject cohen’s dz of 2.2 for brain fluid flow differences between sleep and wakefulness, and 0.9 for the effect of adrenergic inhibition. With an alpha level of 0.01 and 85% power, the estimated sample sizes were *N* = 7 and *N* = 20 for the two parts, respectively. Thus, a study cohort of *N* = 20 (10 in each randomisation sequence) was chosen.

### Study population for sleep study

Twenty healthy males, aged 18-29 years, were included in and completed the sleep study (see flow diagram in Fig S6). Participants were recruited from a local database of individuals who had expressed interest in participating in brain imaging studies and via a national test subject recruitment database (‘www.forsøgsperson.dk’). All participants were right-handed, fluent in Danish, had normal blood pressure, reported no sleep issues when questioned systemically and had no prior or current neurological or psychiatric disorders, learning disabilities, severe somatic diseases, or any use of prescription drugs of relevance for the study. All were non-smokers and did not have excessive use of alcohol or illicit drugs and had not performed shiftwork three months prior to study participation. Participants who had crossed two or more time zones were required to have at least 14 days of recovery per time zone before enrolment. Before inclusion, participants underwent a polysomnographic (PSG) screening night to rule out undiagnosed sleep disorders or low sleep efficiency (defined as < 85%).

### MRI acquisition and reconstruction

MRI was performed on a Siemens (Erlangen, DE) MAGNETOM 3T Prisma scanner with a 64-channel head coil. Structural images were acquired using a high-resolution, whole-brain, T1-weighted MPRAGE scan with the following parameters: Inversion time = 900 ms, repetition time = 1900 ms, echo time = 2.52 ms, flip angle = 9°, in-plane matrix = 256 × 256, in-plane resolution = 0.9 × 0.9 mm, slices = 208, slice thickness = 1.0 mm. For functional imaging, we used a Magnetic Resonance Encephalography (MREG) sequence obtained from the University of Freiburg^63,64^. MREG allows for 3D whole-brain imaging with a repetition time of 100 ms at a resolution of 3×3×3 mm using a stack-of-spirals k-space undersampling trajectory. Further scan parameters were: 3D matrix size = 64×64×50, echo time = 33 ms, flip angle = 25°, field of view = 150 mm and a gradient spoiling of either 0.1 (n = 19) or 1.0 (n = 3) ^34^. The superior part of the field of view was aligned to the superior part of the cortex, to ensure that the majority of the cerebrum, including the third and lateral ventricles, were imaged for all individuals. MREG scans were roughly 5 minutes long. Before scan start, participants were fitted with an MRI compatible respiratory belt and a pulse oximeter wired directly to the scanner. They also wore earplugs to reduce scanner noise, and head cushions were used to minimize movement and ensure consistent head position across scans. Using a MATLAB reconstruction tool provided by the sequence developers^65^, MREG data were reconstructed using L2-Tikhonov regularization with lambda = 0.2. The regularization parameter was determined by the L-curve method. In addition to MREG, 5-min multiband echo-planar images were also acquired after approximately every second MREG scan in the sleep study. However, these were not analysed further in this paper.

### EEG and ECG acquisitions and artefact rejection during MRI

To monitor sleep during MR imaging, participants wore an MRI compatible EEG cap (Electrical Geodesics, Inc., Eugene, OR) with 256 EEG channels and a single reference electrode (Cz) during all scan sessions. The cap was selected according to head circumference of the individual and prepared with an electrolyte/shampoo solution. Cap placement was aided by placing the reference electrode at the vertex and visual inspection of predetermined reference points. The cap was kept in place by a net and kept moist by a shower cap. Impedances were kept below 50 kΩ. Two electrocardiographic (ECG) electrodes were placed on the left side of the sternum, one at the 3^rd^ intercostal space and one at the 6th intercostal space. Inside the head coil, optical cables from the cap were run along the side of the head downwards towards the feet. EEG and ECG data were acquired with a 1 kHz sampling rate and all cables were connected to a synchronization box to match acquisition to the MR scanner clock frequency. The removal of gradient and ballistocardiographic artefacts from the EEG was performed with MATLAB (R2014a, The MathWorks, Inc.) using standard methods in a stepwise approach (see supplements).

### Sleep scoring

All EEG recordings were visually scored in 30 second epochs in accordance with standard AASM criteria (American Academy of Sleep Medicine, 2007) using DOMINO scoring software (Somnomedics, Germany). For overnight and prolonged wakefulness EEG-recordings, sleep was scored by a single expert blinded to the treatment condition, taking the artefact rejection and filter setting into consideration (see Supplementary Materials for details on EEG-acquisition outside of MR). To accurately quantify sleep in EEG data acquired during MR imaging, recordings were scored by two independent experts blinded to subject, treatment and scan type (lights on or lights off). The experts were further instructed to mark epochs with residual MR artefacts and aberrant non-physiological data as “artefacts”. The overall agreement between the two scorers was 73%, similar to what would be expected for standard polysomnography recordings in a non-MR environment^66^.

### Quantitative analyses of EEG during MRI

For quantitative analysis of EEG, data were cleaned subject- and condition-wise by rejecting channels exceeding a kurtosis threshold of 5 using EEGlab’s pop_rejchan function^67^. The leads C3-M2 and C4-M1 were constructed either from preselected channels (Chan_{C3}_ = 59, Chan_{C4}_ = 183, Chan_{M1}_ = 94, Chan_{M2}_ = 190) or, if these were rejected, an average of the neighbouring channels. Artefact segments were rejected from the two leads in 0.5s windows with 0.25s overlap if the power in the 35-120Hz range exceeded 15dB. Delta power ratio was quantified epoch-wise in the two leads using MATLAB’s bandpower function on epoch data, where artefact segments had been rejected. Bandpower (mV^2^) was computed in the 1– 4 Hz range and in the 0.5 – 30 Hz range, where relative delta-power was defined as the ratio of the two. The reported delta power ratio is a mean of the two leads. 30-sec epochs classified as “artefact” by either of the two EEG-scorers were excluded from analysis of EEG delta power ratio.

### MREG and structural brain data preprocessing

The MREG data were stored and reconstructed as detailed above. To ensure steady-state signal saturation, 10 seconds (= 100 images) of data were excluded from the beginning of all MREG scans. Next, MREG data were preprocessed and analysed using Statistical Parametric Mapping software (SPM12, Welcome Trust Center for Neuroimaging, UCL) in MATLAB (R2017b, The Mathworks, Inc.), including motion correction, which was performed by realignment and co-registration of the high-resolution T1 structural image to the MREG data. Co-registration was enhanced with FSL’s Brain Extraction Tool, employing a fractional intensity threshold of 0.3 on the first functional volume of each MREG session, to which the structural image was co-registered. Subsequently, the co-registered structural images were segmented into grey matter, white matter, and cerebrospinal fluid (CSF) maps. In order to evaluate CSF oscillations with as little interference of surrounding tissue and vascular pulsations as possibly, we created a CSF ROI in the left lateral ventricle: We marked a sphere with a radius of 2 cm around a manually selected midpoint of the left ventricle defined in MNI space (MNI coordinates: (8,-10,24)), keeping only the voxels with a segmentation-based CSF probability larger than 0.5. The ventricle ROI were then normalized to each scan session and used to asses CSF oscillations in tissue-type analyses.

### MREG data analysis

For analysis of LFO brain pulsations, we used the entirety of the 5-min MREG sequences and assessed spectral power in a frequency interval defined based on the clinically described B-waves (0.5–2 waves pr. min; 0.008 – 0.033 Hz)^68^. Data from all 5-min MREG scans, corresponding to 10 scored EEG epochs, were high-pass filtered with a 10-order IIR filter (highpass 0.008 Hz). For each 5-min scan, spectral analysis was performed for individual voxels in a whole-brain mask consisting of all voxels with either a grey matter, white matter, or CSF probability larger than 0.1, using the MATLAB periodogram function with a Hanning window and bin width of 0.00244 Hz. The voxel-wise MREG spectra were then averaged to create respective whole-brain or tissue-type-specific MREG power spectra. 5-min scans with framewise displacement >3 mm in one or more epochs were excluded from analysis. To avoid spill-over effects from the bandpass filtering, the exact frequency interval for LFO analysis was set to 0.01221 – 0.03418 Hz (∼ 0.7–2.1 waves pr. min). MREG data were selected for analysis based on the EEG-scoring from two independent experts. Each 5-min scan was included in LFO-analysis as either wakefulness or NREM sleep if at least 80% (8/10 epochs) of the simultaneously recorded EEG epochs were classified as such. 30-sec EEG epochs were labelled as ‘wakefulness’ if they occurred before the first onset of NREM sleep stages N2 or N3 and if both raters either agreed on ‘wakefulness’ or if one classified the epoch as ‘wakefulness’ and the other classified it as ‘artefact’. Epochs were included as ‘NREM sleep’, if both raters classified them as either N2 or N3 NREM sleep. For the 5-min dataset, we only included wakefulness data from lights-on scans and sleep data from lights-off scans (Table S5).

For analyses of respiration- and cardiac-related brain pulsations, we used a 30-sec dataset and assessed spectral power in individually tailored respiration and cardiac frequency bands. Data from all 5-min MREG scans were high-pass filtered with a 10-order IIR filter (passband 0.1 Hz), and were segmented into 30-sec epochs, which were temporally aligned with corresponding sleep-scored EEG epochs. For each 30-sec epoch, spectral analysis was performed as described for the 5-min dataset, but with a bin width of 0.02 Hz. Epochs with a framewise displacement above 3 mm were excluded (n = 91 epochs). We defined epoch-by-epoch heart rate and respiration frequencies using temporally aligned data from the respiratory belt and the pulse oximeter, imported using the TAPAS PhysIO toolbox^69^. The dominant peaks in respiration (0.13 – 0.5 Hz; 7.8 – 30 min^-1^) and cardiac frequency ranges (0.67 – 2 Hz; 40.2 – 120 min^-1^) were determined from the periodogram-determined power spectrum of each 30-sec epoch of physiological data. The epoch-by-epoch peak frequencies for respiration and heart rates were then used to define the individually tailored 30-sec epoch-specific respiration and cardiac frequency bands in the MREG spectra. The width of the respiration bands was defined as the epoch-wise mean respiration rate ± 0.06 Hz (± 3 bins), and the width of the cardiac band was defined as the epoch-wise mean heart rate ± 0.1 Hz (± 5 bins). 30-sec epochs with non-physiological values (< 40 or > 90 heart beats per min. and/or < 8 or > 25 breaths per min.) were excluded. All 30-sec MREG epochs labelled as wakefulness or NREM sleep were included in analyses assessing effects of sleep and sleep deprivation on spectral power in respiration and cardiac frequency bands (Table S3).

For sleep-depth analyses, epochs were only included as NREM sleep stage N2 or N3 if there was a consensus between raters on the specific sleep stage (Table S6). In analyses of the relationship between EEG delta power ratio and spectral power, all 30-sec epochs classified as “artifact” by any of the scorers were excluded. All other epochs were included, regardless of scored vigilance state.

### Psychomotor vigilance

A psychomotor vigilance test (PVT; e-Prime software, Psychology Software Tools Inc., Pittsburgh) was used to assess psychomotor vigilance cognitive effects of prolonged wakefulness^70^. PVT is a simple reaction time task where participants press the space key as quickly as possible upon seeing a digital millisecond counter on the computer screen. For each PVT trial, 100 stimuli were presented (random interstimulus intervals: 2–10 sec). Participants completed the PVT both before scans (rested: 7 hr awake; sleep-deprived: 31 hr awake) and immediately after exiting the scanner (1 hr sleep opportunity) (Fig 2). Participants completed a training session in their adaptation week. Two validated PVT variables were quantified^50,51^: “median reaction time” and “lapses of attention (reaction time > 500 ms). Impaired psychomotor vigilance due to sleep deprivation was evaluated by 1) quantifying the increase in median reaction time from rested wakefulness (measure before sleep-deprived scan – measure before well-rested scan) and 2) the absolute number of lapses of attention before the sleep-deprived scan (Freeman-Tukey transformed: 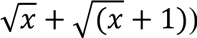. Improvements in psychomotor vigilance due to sleep in the scanner were computed as the difference between measures taken before and immediately after scans.

### Valsalva Manoeuvre and breath-holding trial

To validate our interpretation of MREG-detected brain oscillations, we conducted a descriptive study assessing the effects of breath-holding and the Valsalva manoeuvre on the MREG-signal. We included four healthy participants aged 31-40 years (two males) without prior or current major medical or psychiatric diagnoses, and no use of psychoactive medications. A single scan session was conducted between 4 and 6 PM. The following three manoeuvres (1 minute break in between) were repeated three times while the MREG sequence ran continuously: 30 sec rest, 30 sec breath-hold and 30 sec Valsalva manoeuvre. Participants were instructed in and practiced the manoeuvres just prior to scan start, with specific instructions to avoid inhaling before breath-holding to prevent inducing a low-pressure Valsalva manoeuvre. While in the scanner, instructions and timings were displayed on a monitor visible to the participants. To ensure and monitor wakefulness, lights were kept on and participants were encouraged to keep their eyes open.

Scan setup, scan parameters and reconstruction of the MREG data were identical to the sleep study (as described above). MREG-scans were divided into 25-30 sec epochs matching the three conditions (rest, breath-hold or Valsalva manoeuvre) and high-pass filtered with a 10-order IIR filter (passband 0.02 Hz). Data-frames with motion artefacts related to initiation of the Valsalva manoeuvre or breath-holding were discarded, and scans with bigger artefact during the manoeuvres were excluded. Epoch-wise spectral analysis was performed as described above for the 30-sec dataset in the sleep study. To investigate effects of the manoeuvres on the MREG-signal, we evaluated power peaks in whole-brain MREG-spectra within LFO (0.02 – 0.1 Hz), respiration (0.14 – 0.5 Hz) and cardiac (0.68 – 2 Hz) frequency ranges. Because 30-sec time series are too short for evaluation of spectral power in the LFO band applied in the sleep study (∼ 0.7 – 2 waves per min.; 0.012 – 0.034 Hz), we restricted our interpretations of spectral power intensity in low frequency ranges to only conclude on what happens at or below 0.1 Hz in the MREG spectra.

### Statistical analyses

Whole-brain or regional MREG spectral power within LFO, respiration and cardiac frequency bands (sum of spectral power within frequency range) from well-rested scan sessions (awake scan), sleep deprived placebo scan sessions (awake and sleep scan) and sleep deprived carvedilol scan sessions (awake and sleep scan) were log transformed to mitigate skewness in the distribution before being analysed using linear mixed models. Sleep deprivation (well-rested vs sleep-deprived), vigilance state (awake vs deep NREM sleep), sleep depth (sleep-deprived awake vs N2 vs N3), tissue type (grey matter vs white matter vs CSF), treatment (placebo vs carvedilol; randomised to sleep deprived scan session 1 and 2), PVT measurements (‘median reaction time‘ and ‘lapses of attention’) and delta power ratio were included as additive fixed effects, with interactions between vigilance state and tissue type as well as between vigilance state and treatment. Subject ID, scan session (well-rested, sleep-deprived session 1, sleep-deprived session 2) and sleep opportunity (lights on, lights off) were included as random intercepts with sleep opportunity being nested into scan session itself nested into subject ID. To account for the impact of declining spectral power as a function of higher frequencies^71^, respiration or heart rates recorded simultaneously with MREG-data were also included as fixed effects in all models assessing spectral power within respiration and cardiac frequency bands. P-values for testing fixed effects were obtained using Wald tests. The expected log spectral power at various conditions (e.g. sleep-deprived under carvedilol) was computed from the mixed model estimates to illustrate the model fit.

Associations between (*i*) frequencies of measured respiration/heart rates and corresponding peaks in the MREG spectra, (*ii*) PVT measurements and mean log spectral power within LFO, respiration and cardiac frequency bands in 5-min scans/30-sec epochs and (*iii*) delta power ratio and log spectral power in respiration and cardiac frequency bands in 30sec epochs were assessed with Wald tests obtained from linear mixed models (denoted *p*_adj_). Corresponding correlations coefficients were deduced from mixed model estimates (denoted *r*_adj_). For reference, Pearson’s correlation coefficients for associations between *(i)*, *(ii)* and *(iii)* were also evaluated (regardless of subject ID, scan session, sleep opportunity, respiration- and heart rates and treatment) and denoted *r*_raw_.

Where pairwise testing was appropriate instead of linear mixed models, we conducted Student’s two-tailed t-tests for data with a normal distribution and Wilcoxon signed-rank tests for data not normally distributed.

P-values equal to or below 0.05 were considered significant except in analyses where three levels of a fixed effect were evaluated (and compared pairwise). Here, P-values were adjusted for multiple comparison with Bonferroni correction. All analyses were performed with the statistical software R (http://www.R-project.org/). The R packages lme4 (version: 1.1.29), lmerTest (version: 3.1.3), and LMMstar (version: 0.8.9) were used for, respectively, fitting random intercept models, performing Wald tests, and estimating expected means and correlation parameters.

For detailed descriptions of all statistical models applied throughout the paper, see ‘*statistical models for all reported results*’ in Supplementary Materials.

### Study approvals

The studies were approved by the Scientific Ethics Committees of the Capital Region of Denmark (H-16045933 & H-18048865). They adhered to the principles of the Declaration of Helsinki. Written informed consent was obtained from all participants before enrolment in the studies.

## Acknowledgments

We would like to thank Helle Leonthin for sleep staging and Emily Beaman, Cecilie Lerche Nordberg as well as members of the Center for Translational Neuromedicine lab for their assistance with data collection and supervision of proper sleep deprivation. We also thank D. Xue from the Center for Translational Neuromedicine for assistance with illustrations.

The research reported in this work was funded by the H2020 Marie Skłodowska-Curie Actions grant: H2020-MSCA-IF-2017-798131 (SCH); the Lundbeck Foundation grant: R264-2017-2778 (SCH); the Lundbeck Foundation grant: R279-2018-1145 (GMK); the Independent Research Fund Denmark grant: 0134-00454B (GMK); the Independent Research Fund Denmark grant: 7025-00090B (GMK); Rigshospitalet Forskningspuljer grant: R151-A6534 (GMK); the EU Joint Programme - Neurodegenerative Disease Research 2022-120 (VK, GMK, MN). The funders had no role in research conceptualization, study design, data collection and analyses, decision to publish or manuscript preparation.

## Author contributions

GMK, SCH, MN and PJJ conceptualised and designed the study. SCH and SMUL set up the sleep study protocol. SCH, SMUL, DBZ and SP were responsible for data collection. SMUL conducted analyses of sleep study data with the help of SCH, ASO, DBZ and KBB. DBZ scored EEG-data. ASO preprocessed EEG- and neuroimaging data from the sleep study and evaluated voxel-wise spectral power. PW measured norepinephrine levels in serum from sleep study. SMUL and ASO conducted the Valsalva- and breath-holding trial and analysed this data. BO set up statistical models and helped interpret model outcomes and visualise results. VK and PJJ gave technical support and conceptual advice on EEG and MREG-data collection and analysis. GMK, MN and PJJ supervised the project. SMUL, SCH and GMK wrote and prepared the manuscript. All authors contributed to the interpretation of result and have all approved the final version of the manuscript.

## Competing interests

S.C.H. is currently an employee of Roche Pharma, which is unrelated to the contents of this manuscript. All other authors declare no competing interests.

## Data and code availability

All data and code underlying the findings of this study will be available from the corresponding author upon reasonable request.

## List of Supplementary Materials

Supplementary figures (S1 to S6)

Supplementary tables (S1 to S6)

Supplementary methods

Supplementary references

## Notes

### Clinical Trial

Clinical trial ID NCT03576664

### Author Declarations

Scientific Ethics Committees of the Capital Region of Denmark gave ethical approval for this work

