## Supplementary Materials for "Sleep pressure propels cerebrovascular oscillations while sleep intensity correlates with respiration- and cardiac-driven brain pulsations"

**This PDF file includes:**

Supplementary figures (S1 to S6)
Supplementary tables (S1 to S6)
Supplementary methods
Supplementary references[1–4]

### SUPPLEMENTARY FIGURES

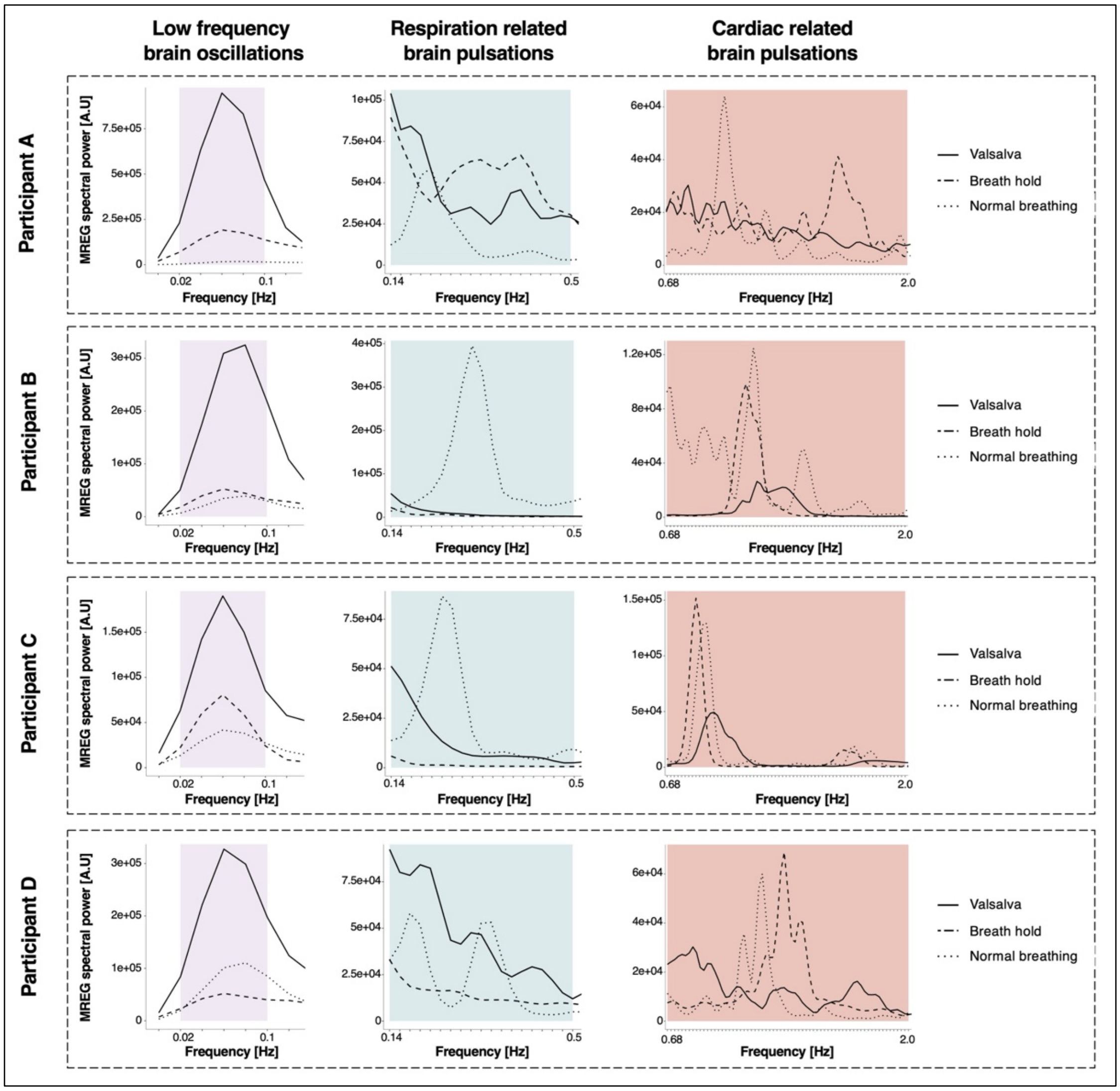


**Fig. S1. Effect of physiological modulations of intracranial pressure, respiration and cardiac action on MREG spectral power for all 4 included individuals.** Participant-wise (participant A-D) mean whole-brain MREG spectra during normal breathing, breath-holding and the Valsalva manoeuvre, averaged across 3 imaging sessions. Shaded areas represent frequency ranges in which spectral power and power peaks are evaluated. Of note, participant A and D exhibit heightened spectral power and a series of smaller peaks in the respiration band (blue shading) during both the Valsalva Manouvre and breath-holding, besides the expected respiration-related peaks during normal breathing. We interpretate these additional peaks in power as harmonics from the strong power peaks in the LFO band (purple shading).

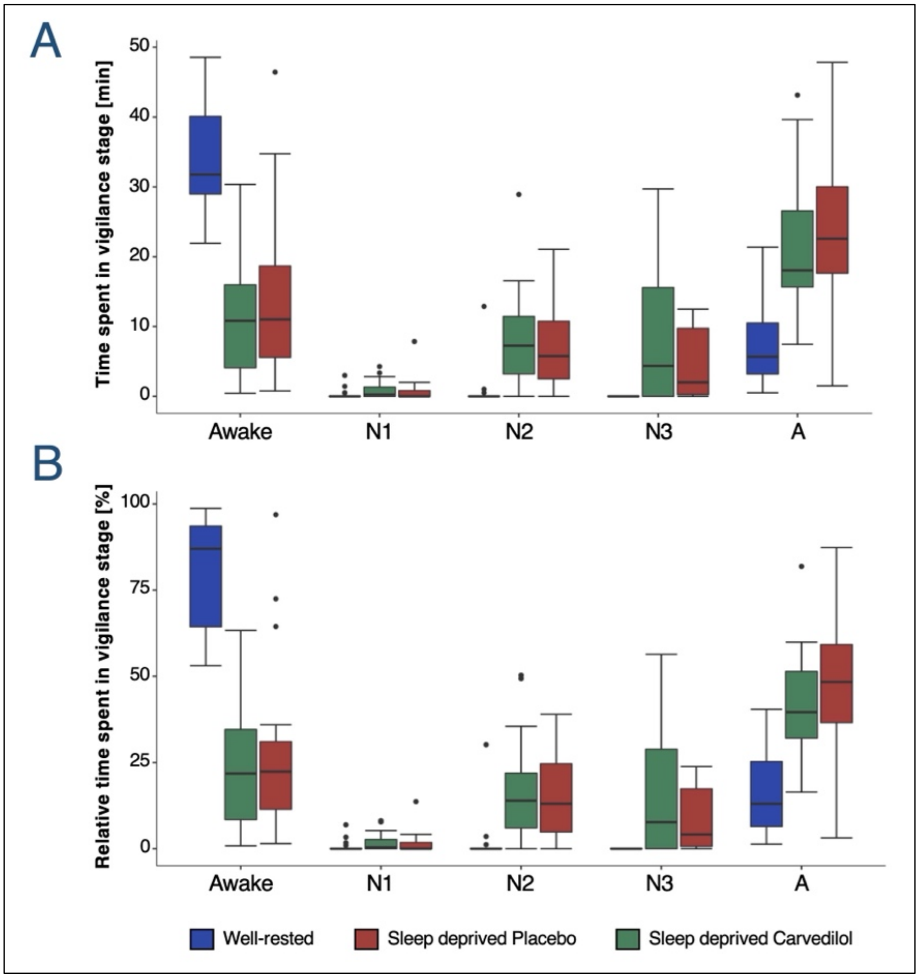


**Fig. S2. EEG-recorded vigilance states during MR-EEG sessions.** Boxplots illustrate absolute **(A)** and relative **(B)** time spent in wakefulness and NREM sleep stages N1, N2 and N3 during rested and sleep-deprived (placebo and carvedilol) scan sessions. All 30-sec EEG epochs recorded simultaneously with MR-scans were included in analyses. EEG-epochs were included in analyses when the two independent EEG scorers agreed on staging. Epochs with scorer disagreement and epochs scored as artifacts were categorized as ‘A’. There is no difference between carvedilol and placebo conditions (paired t-test, $p_{all} > 0.05$). Box-plot elements include: median (center line), upper and lower quartiles (box limits), 1.5x interquartile range (whiskers) and outliers (points). $N = 20$.

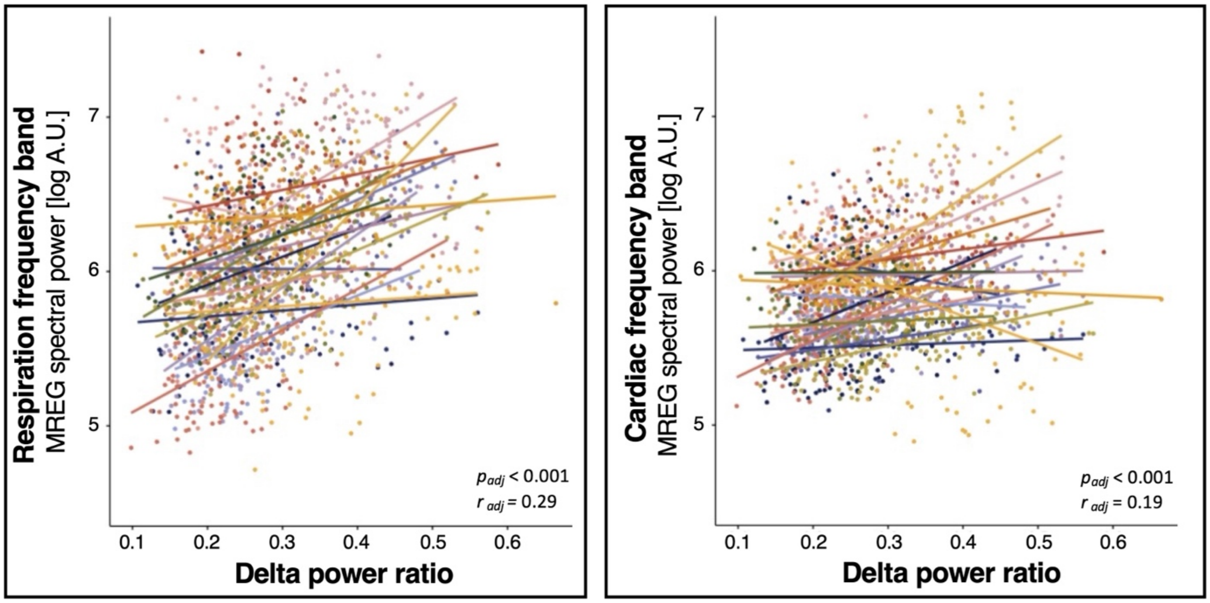


**Fig. S3. Participant-wise correlations between EEG delta power ratio as recorded simultaneously with MREG-imaging, and spectral power.** Each dot represents a 30 sec epoch and each participant's data is shown with a unique colour. $r_{est}$ & $p_{est}$ are estimated correlation coefficient and estimated p-value from linear mixed models, where repeated measurements are taken into account. **(A)** A positive slope was observed for 17 of 19 participants in the respiration frequency band, and of these 13 were statistically significantly positively correlated. **(B)** For the cardiac band, 16 of 20 participants were positive and 11/20 had a significant positive slope.

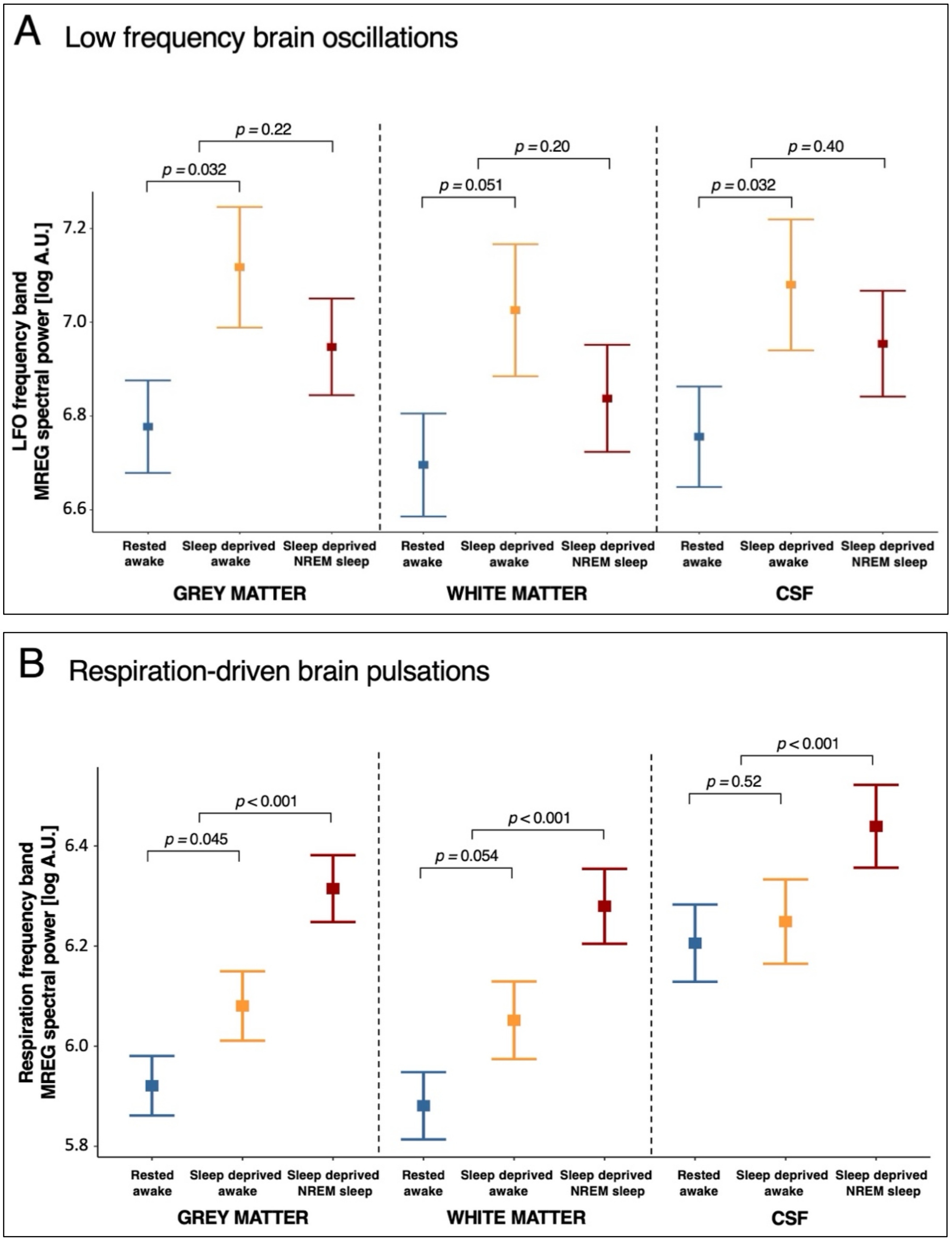

A Low frequency brain oscillations
p = 0.22
p = 0.032
p = 0.20
p = 0.051
p = 0.40
p = 0.032
LFO frequency band
MREG spectral power [log A.U.]
7.2
7.0
6.8
6.6
Rested awake
Sleep deprived awake
Sleep deprived NREM sleep
GREY MATTER
WHITE MATTER
CSF
B Respiration-driven brain pulsations
p < 0.001
p = 0.045
p < 0.001
p = 0.054
p < 0.001
p = 0.52
Respiration frequency band
MREG spectral power [log A.U.]
6.4
6.2
6.0
5.8

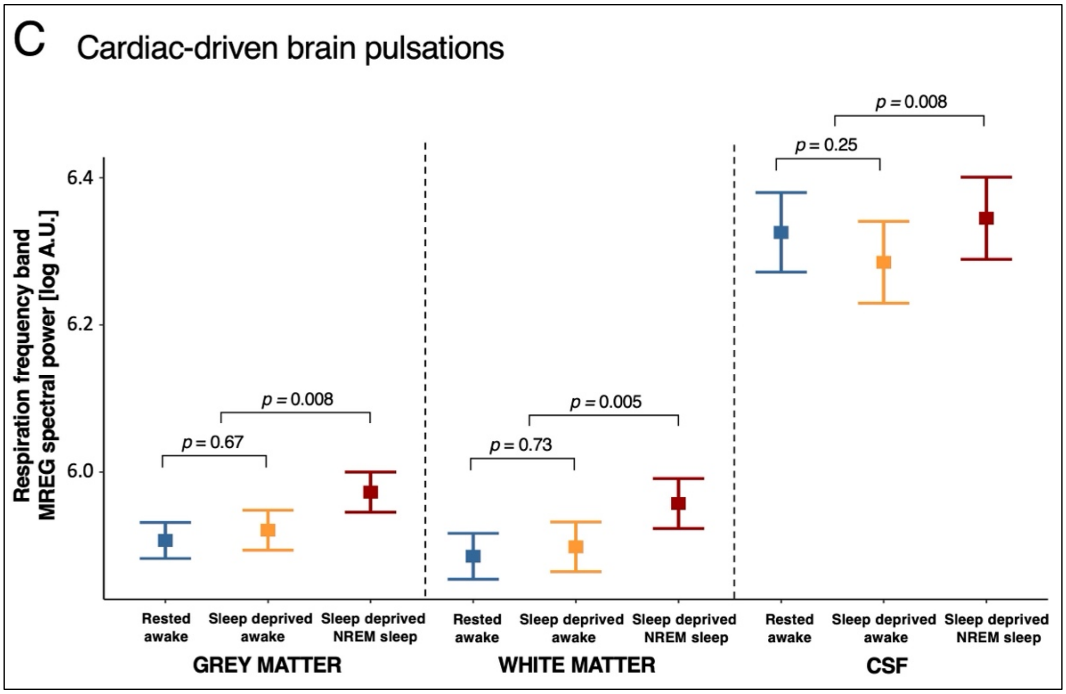


**Fig. S4. Effects of sleep deprivation and NREM sleep (combined N2 and N3) on brain oscillations across grey matter, white matter and CSF.** Results from sensitivity analyses of the effects of sleep deprivation and NREM sleep on MREG power in the **(A)** LFO, **(B)** respiration, and **(C)** cardiac frequency bands across three tissue types (grey matter, white matter and CSF). Error plots represent estimates (estimated means ± SEM) from linear mixed models, run separately for each tissue type.

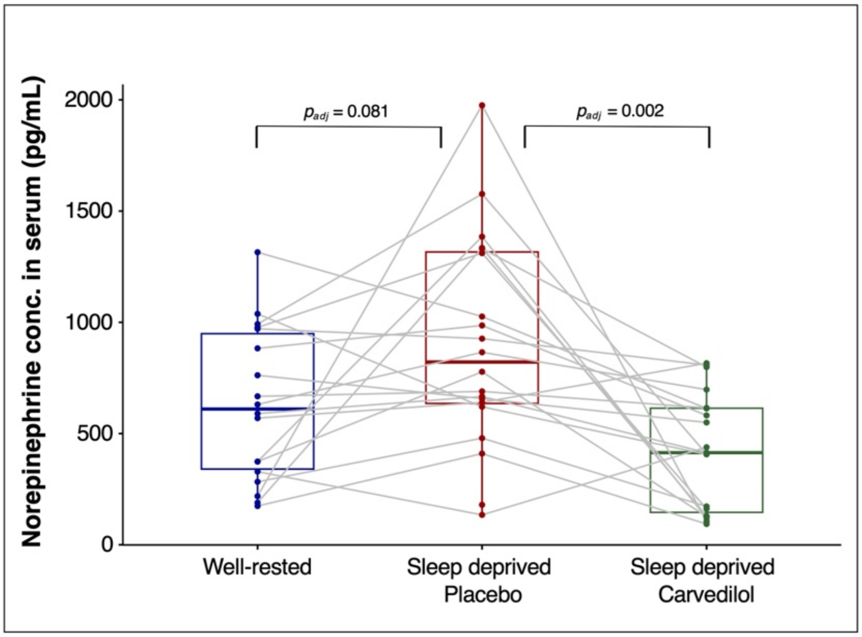


**Fig. S5. Serum norepinephrine (pg/mL) immediately after all scans.** Normal range: 200-1700 pg/mL. p-values are from paired student's t-tests and have been adjusted for multiple comparisons with Bonferroni corrections. Box-plot elements include: median (center line), upper and lower quartiles (box limits) and 1.5x interquartile range (whiskers). Well-rested vs sleep- deprived placebo: $N = 18$. Sleep-deprived placebo vs carvedilol: $N = 19$.

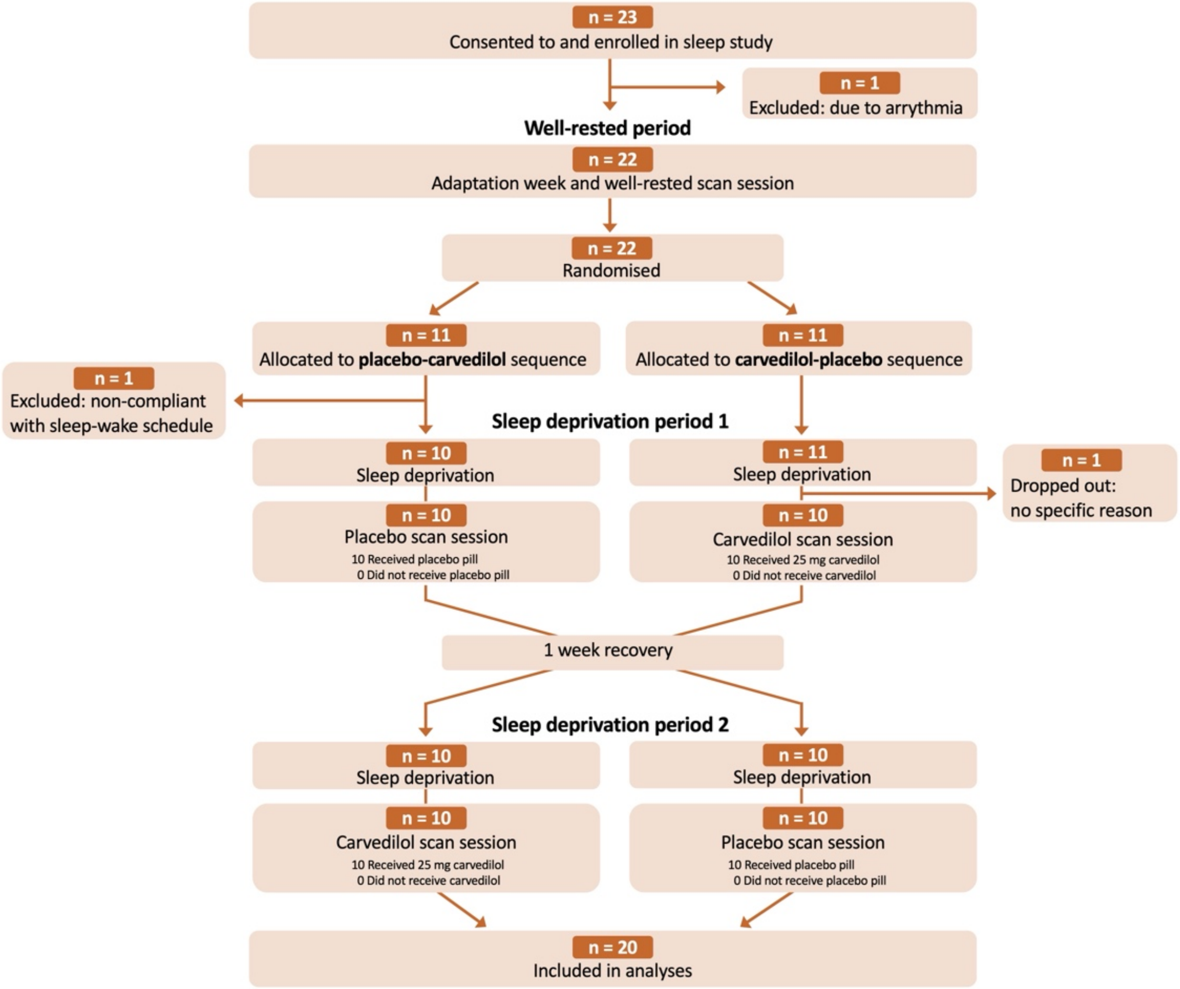


**Fig. S6.** Consort flow diagram presenting number of research volunteers enrolled, excluded, allocated to intervention sequence and who completed the study and were included in analyses.

### SUPPLEMENTARY TABLES

#### Table S1. Wakefulness before MR-scan sessions.

| | Before well-rested scan | Before sleep-deprived scan PLACEBO | Before sleep deprived scan CARVEDILOL | *Diff. in wakefulness before placebo and carvedilol scans* |
|---|---|---|---|---|
| **Hours from wake-up time to MR-scans (h)** | 11.1 ± 0.4 | 34.9 ± 0.3 | 34.8 ± 0.3 | *p* = 0.21 |
| **Duration of prolonged wakefulness EEG-recording (h)** | - | 29.7 ± 3.2 | 28.5 ± 5.8 | *p* = 0.33 |
| **NREM sleep (min)** | - | 1.4 ± 5.2 | 0.8 ± 1.7 | *p* = 0.56 |
| **REM sleep (min)** | - | 0.0 ± 0.0 | 0.0 ± 0.0 | *NA* |

Time participants spent awake between waking from their 8-hours standardised sleep to the beginning of scan sessions. *p*-values represent results from paired t-tests. $N = 20$, all data are shown as mean ± SD. NREM sleep: Non-rapid eye movement sleep stages N1-N3, REM sleep: Rapid eye movement sleep.

**Table S2. Data across the three EEG-monitored nights of 8-hr standardised sleep leading up to the MR imaging sessions**

| | Night before well-rested | Night before sleep-deprived PLACEBO | Nights before sleep-deprived CARVEDILOL | *Diff. between nights* |
|---|---|---|---|---|
| **TIB (h)** | 7.9 ± 0.2 | 7.9 ± 0.3 | 7.9 ± 0.1 | *p = 0.34* |
| **TST (h)** | 7.4 + 0.3 | 7.4 ± 0.3 | 7.4 ± 0.3 | *p = 0.16* |
| **Sleep efficiency (%)** | 93.6 ± 3.4 | 94.4 ± 2.7 | 93.8 ± 3.5 | *p = 0.25* |
| **NREM sleep (h)** | 5.5 ± 0.3 | 5.4 ± 0.4 | 5.4 ± 0.4 | *p = 0.21* |

Estimates and *p*-values are from a linear mixed model (hours of NREM sleep) and Friedman tests (TIB, TST & Sleep efficiency). all data are shown as mean ± SD. *N* = 20. TIB: time in bed, TST: Total sleep time, Sleep efficiency: % time spent asleep of time spent in bed (after lights are off), NREM sleep: duration of NREM sleep stages N1, N2 and N3.

**Table S3. Data included in 30-sec dataset for analyses of sleep deprivation- and NREM sleep effects on spectral power in respiration and cardiac frequency bands.**

| | | PLACEBO | | CARVEDILOL | |
|---|---|---|---|---|---|
| | Well-rested wakefulness | Sleep deprived wakefulness | Sleep deprived deep sleep (N2N3) | Sleep deprived wakefulness | Sleep deprived deep sleep (N2N3) |
| Participants included in analysis (N) | 20 | 17 | 17 | 15 | 19 |
| Included 30-sec MREG epochs (n) | 35.6 [28.1, 43.0] | 10.8 [5.2, 16.3] | 28.7 [20.2, 37.2] | 9.5 [4.7, 14.4] | 33.8 [25.6, 42.1] |
| Respiration rate ($min^{-1}$) | 15.2 [14.1, 16.4] | 16.2 [14.9, 17.7] | 14.9 [13.7, 16.2] | 16.2 [14.8, 17.7] | 14.3 [13.2, 15.5] |
| Heart rate ($min^{-1}$) | 60.9 [57.5, 64.4] | 59.5 [56.1, 63.1] | 53.8 [50.8, 57.1] | 59.6 [56.2, 63.2] | 54.8 [51.7, 58.0] |
| Mean arerial pressure (mmHg) | 91.3 [89.4, 93.3] | 92.5 [90.1 – 94.9] | | 88.2 [85.8, 90.5] | |
| Pulse pressure (mmHg) | 45.7 [42.4, 48.9] | 45.2 [42.1, 48.2] | | 44.3 [40.7, 47.9] | |

Values are shown as mean and 95% confidence intervals and are determined from linear mixed models to account for interindividual variance. Participants included in analysis: Number of participants in each condition, who had at least one 30-sec MREG-epoch with EEG-confirmed vigilance state (wakefulness or NREM sleep stage N2+N3; with agreement between the two sleep scoring experts). Respiration and heart rates: Physiological measurements recorded simultaneously with MREG. Respiration and heart rates are estimated for all 30 sec MREG-epochs and used to determine epoch-wise respiration and cardiac frequency bands. Mean arterial blood pressure (1/3*diastolic blood pressure) + (2/3*systolic blood pressure) and pulse pressure (difference between systolic and diastolic blood pressure): Average of measures performed immediately before and after MR-scans.

**Table S4. Recovery sleep characteristics after sleep deprivation in each treatment arm.**

| | RECOVERY NIGHTS | | |
|---|---|---|---|
| | **PLACEBO** | **CARVEDILOL** | **Treatment P-value** |
| **TIB (h)** | 8.0 ± 0.0 | 7.9 ± 0.3 | $p = 0.37$ |
| **TST (h)** | 7.8 ± 0.1 | 7.7 ± 0.4 | $p = 0.39$ |
| **Sleep efficiency (%)** | 97.3 ± 1.5 | 97.1 ± 1.6 | $p = 0.60$ |
| **Sleep latency (h)** | 0.1 ± 0.1 | 0.1 ± 0.1 | $p = 0.71$ |
| **REM latency (h)** | 1.4 ± 0.8 | 1.2 ± 0.7 | $p = 0.27$ |
| **NREM total (%)** | 70.6 ± 3.8 | 70.9 ± 3.5 | $p = 0.51$ |
| **REM total (%)** | 26.8 ± 3.9 | 26.3 ± 3.6 | $p = 0.38$ |
| **Stage N1 (%)** | 3.5 ± 2.3 | 2.6 ± 1.2 | $p = 0.03$ |
| **Stage N2 (%)** | 37.4 ± 5.9 | 39.4 ± 7.0 | $p = 0.10$ |
| **Stage N3 (%)** | 29.7 ± 7.2 | 28.9 ± 8.4 | $p = 0.76$ |
| **WASO (%)** | 1.4 ± 0.9 | 1.7 ± 1.2 | $p = 0.31$ |

Sleep stages are presented as a percentage of total time in bed. Analyses of the recovery nights were restricted to the first 8 hours (480 minutes). *p*-values are from paired students t-tests (REM latency, NREM total, REM total, Stage N2, stage N3) and Wilcoxon signed rank tests (TIB, TST, Sleep efficiency, Sleep latency, Stage N1). $N = 20$, all data are shown as mean ± SD. TIB: time in bed, TST: total sleep time, Sleep latency and REM latency: time from lights-off to the first occurrence of stage N2 sleep and REM sleep, NREM: non rapid eye movement sleep, REM: Rapid eye movement sleep WASO: wakefulness after sleep onset.

**Table S5. Data included in 5-min dataset for analyses of sleep deprivation and NREM sleep effects on spectral power in LFO (0.012 - 0.034 Hz) frequency band.**

| | | PLACEBO | | CARVEDILOL | |
|---|---|---|---|---|---|
| | Well-rested wakefulness | Sleep deprived wakefulness | Sleep deprived deep sleep (N2N3) | Sleep deprived wakefulness | Sleep deprived deep sleep (N2N3) |
| **Participants included in analysis (N)** | 19 | 12 | 14 | 12 | 17 |
| **Included 5min MREG scans (n)** | 1.6 [1.3, 1.8] | 1.1 [0.9, 1.3] | 3.1 [2.1, 4.1] | 1.3 [0.7, 1.8] | 3.3 [2.5, 4.1] |

Values are shown as mean and 95% confidence intervals and are determined from linear mixed models to account for interindividual variance. Participants included in analysis: Number of participants in each condition, who had at least one 5-min MREG scan with 80% (8/10 epochs) EEG-confirmed vigilance state (see methods).

**Table S6. Data included in 30-sec data set for sleep depth analyses.**

| | PLACEBO | | CARVEDILOL | |
|---|---|---|---|---|
| | N2 sleep | N3 sleep | N2 sleep | N3 sleep |
| **Participants included in analysis (N)** | N = 16 | N = 14 | N = 17 | N = 12 |
| **Included 30-sec MREG epochs (n)** | 14.5 [9.1 – 19.9] | 8.1 [4.5 – 11.7] | 14.6 [8.9 – 20.3] | 17.8 [7.1 – 28.6] |
| **Respiration rate ($min^{-1}$)** | 14.5 [13.4 – 15.6] | 14.4 [13.3 – 15.6] | 14.2 [13.4 – 15.3] | 13.6 [12.6 – 14.7] |
| **Heart rate ($min^{-1}$)** | 52.3 [49.5 – 55.2] | 53.9 [51.0 – 56.9] | 54.6 [51.8 – 57.6] | 54.5 [51.6 – 57.5] |

Values are shown as mean and 95% confidence intervals and are determined from linear mixed models to account for interindividual variance. Participants included in analysis: Number of participants in each condition who had at least one 30-sec epoch, where EEG-scorers agreed on either N2 or N3 NREM sleep. Respiration and heart rates were recorded simultaneously with MREG and estimated for all 30-sec MREG-epochs and subsequently used to determine epoch-wise respiration and cardiac

### SUPPLEMENTARY METHODS

Polysomnography and prolonged wakefulness EEG recordings

All sleep and wake EEGs recorded outside the MR environment were recorded using the battery powered and transportable SomnoScreen Plus system (Somnomedics, Germany). Impedance values were kept below 6kΩ at the beginning of all recordings.

Overnight polysomnography recordings were performed in private rooms with blinded windows. Data across 18 EEG electrodes, placed according to the internationally standardized 10–20 system[1], were recorded simultaneously with submental electromyogram, electrooculogram (EOG) and ECG. In order to define the overnight sleep periods, participants were asked to blink ten times when they went to bed and turned off the light and again when they woke up and turned on the light. Participants were prescribed a fixed 'lights on' and 'lights off' time of an 8-hour (standard night) or 10-hour (recovery night) sleep opportunity and data analysis was restricted to a maximum of eight hours (480 minutes) to enable comparisons between nights.

To monitor vigilance during the two periods of sleep deprivation, participants were fitted with a minimal EEG setup with 6 EEG electrodes and EOG. These EEG data were recorded continuously throughout the wakefulness periods. When the EEG equipment was fitted, the recorder was placed in a bag, which allowed participants to move around freely.

Removal of MR-induced artifacts on EEG

First, the Average Artefact Subtraction method was used to eliminate gradient artefacts. This creates a template gradient artefact by using the MR-trigger signal as a time-locking event and then averaging across the nearest 30 artefacts, in a moving window on high-pass filtered data[2].

Second, the Optimal Basis Sets approach was used to adaptively remove ballisto-cardiographic artefacts over time. This approach uses the ECG R-peak as a time-locking event and combines the local moving average template construction with a combination of basis functions, derived from a principal component analysis of the gradient cleaned EEG-signal[3].

Statistical models for all reported results

***(i) Effects of ICP, respiration and cardiac action on MREG spectral power:***

This is a descriptive study ($N$ = 4). Effects of the Valsalva manoeuvre and breath-holding on MREG-detected brain oscillations were evaluated by inspecting how spectral power peaks within LFO, respiration, and cardiac frequency ranges in the MREG-spectra change during the manoeuvres – both on an individual level (Fig S1) and on a MREG spectra averaged across the four participants (Fig 1A-D).

To assess the association between measured respiration and heart rates and the frequency of the corresponding peaks in the MREG spectra, we included all 30 sec epochs recorded during the well-rested scan sessions in the sleep study. We performed a linear mixed model with the frequency of power peaks in respiration/cardiac frequency ranges as the outcome variable, recorded respiration/heart rates as a fixed effect and Subject ID as random intercept. The reported p-values are the ones of the respiration/heart rate parameter in the mixed model (denoted $p_{\text{adj}}$). The corresponding correlation coefficient ($r_{\text{adj}}$) was deduced from the mixed model estimates using an approximation suggested by Lipsitz et al.[4]: $r = \beta / \sqrt{\beta^2 + \text{df}\, \sigma_{\hat{\beta}}^2}$, where $\beta$ denotes respiration/heart rate parameter in the mixed model, $\sigma_{\hat{\beta}}$ its standard error, and $df$ its degree of freedoms. For reference, Pearson correlation coefficients were evaluated (regardless of subject ID) and denoted $r_{\text{raw}}$ (Fig 1E-F). $N_{participants}$ = 20, $N_{epochs}$ = 1148.

***(ii) Sleep study cohort and descriptives:***

Paired two-tailed Student's t-tests were used to compare the mean time intervals from wake-up time to scan start and the mean time spent in NREM/REM sleep during the periods of prolonged wakefulness prior to scans (Table S1), as well as the relative time spent in wakefulness/sleep during MR-scans (Fig S2) between carvedilol and placebo conditions. Paired t-tests were also used to evaluate the difference in reaction time and lapses of attention between rested wakefulness and sleep deprived wakefulness. $N$ = 20.

Sleep during the standardised nights leading up to both well-rested, sleep deprived placebo and sleep deprived carvedilol study sessions were compared by evaluation PSG-recorded sleep patterns (Table S2) between the three nights. A linear mixed model including night type (before-well-rested vs before-placebo vs before-carvedilol) as a fixed effect and subject ID and study week (week 1-3) as random intercepts (study week being nested into subject ID), was used to evaluate mean differences in 'Hours of NREM sleep (h)'. P-value for testing the fixed effect was obtained using a Wald test. Friedman tests were used to evaluate differences in 'Time in bed (h)', 'Total sleep time (h)' and 'sleep efficiency (%)' between the three nights, as these measures were not normally distributed. $N = 20$.

Effect of sleep deprivation and NREM sleep on EEG delta power ratio as well as on respiration- and heart rates (recorded simultaneously with MREG) were assessed in 30sec epochs from well-rested (awake scans) and sleep deprived placebo (awake and sleep scans) conditions (Table S3) with a linear mixed model. Sleep deprivation (well-rested vs. sleep-deprived) and vigilance state (awake vs NREM sleep) were included as additive fixed effects and Subject ID, scan session (well-rested, sleep-deprived scan 1, sleep-deprived scan 2) and sleep opportunity (lights on, lights off) were included as random intercepts with sleep opportunity being nested into scan session itself nested into subject ID. This linear mixed model will be referred to as LMM-*ii*. P-values for testing fixed effects were obtained using Wald tests. $N = 20$.

***(iii) Sleep deprivation enhances spectral power in the LFO frequency band:***

Effects of sleep deprivation and NREM sleep on whole-brain LFOs were evaluated in 5 min MREG scans from well-rested (awake scans) and sleep deprived placebo (awake and NREM sleep scans) conditions. Whole-brain spectral power within the LFO band was calculated by summing the spectral power of all bins included in the 0.01221 – 0.03418 Hz frequency range, after which it was log transformed to mitigate skewness in the distribution before being analysed. A linear mixed model with same fixed and random effect structure as LMM-*ii* was used for analysis and P-values for testing fixed effects were obtained using Wald tests. The expected log spectral power at various conditions (e.g. sleep-deprived wakefulness) was computed from the mixed model estimates to illustrate the model fit (Fig 3B and Fig S4A). $N = 20$ (number of participants with data in each of the three conditions can be seen in Table S5).

The correlation between mean log spectral power in the LFO band (in either sleep deprived awake or NREM sleep scans) and PVT measurements (either pre-scan measurements or difference between pre- and post-scan measurements) was assessed considering two measurements per individual: one from placebo- and one from carvedilol conditions. A Wald test for the association between spectral power and PVT measures was obtained from a linear mixed model based on subject ID and adjusted for treatment and scan session nr (denoted $p_{\text{adj}}$). The corresponding correlation coefficient ($r_{adj}$) was evaluated (without proximation) as follows:

We introduce the following notations:

- $Y_{1,j}$ and $Y_{2,j}$ denote the spectral power at, respectively, placebo and carvedilol for individual $j$.
- $Z_{1,j}$ and $Z_{2,j}$ denote the PVT measurement at, respectively, placebo and carvedilol for individual $j$.
- $X_j$ denotes the covariates relative to individual $j$, here scan week.

We are interested the following correlation coefficient between spectral power and PVT measures:

$$r_{\text{adj}} = cor\left(Y_{1,j}, Z_{1,j}|X\right) = cor\left(Y_{2,j}, Z_{2,j}|X\right)$$

assumed to be independent of the treatment and scan week. We estimate this correlation under the following linear mixed model:

$$Y_{t,j} = \alpha_t + \beta X_{t,j} + u_j + \varepsilon_{t,j}$$

$$Z_{t,j} = \mu_t + \gamma X_{t,j} + v_j + \xi_{t,j}$$

Where $\begin{bmatrix} u_j \\ v_j \end{bmatrix} \sim N\left(\begin{bmatrix} 0 \\ 0 \end{bmatrix}, \begin{bmatrix} \tau_1 & \tau_{12} \\ \tau_{12} & \tau_2 \end{bmatrix}\right)$ and $\begin{bmatrix} \varepsilon_{t,j} \\ \xi_{t,j} \end{bmatrix} \sim N\left(\begin{bmatrix} 0 \\ 0 \end{bmatrix}, \begin{bmatrix} {\sigma_1}^2 & r_{\text{adj}}\sigma_1\sigma_2 \\ r_{\text{adj}}\sigma_1\sigma_2 & {\sigma_2}^2 \end{bmatrix}\right)$

using restricted maximum likelihood (REML).

For reference, Pearson correlation coefficients between mean log LFO spectral power and PVT measures were evaluated (regardless of subject ID, treatment and scan session nr) and denoted $r_{\text{raw}}$ (Fig 4). $N_{awake}$ = 16 and $N_{sleep}$ = 17.

***(iv) NREM sleep enhances MREG spectral power in the respiration and cardiac frequency bands; N3 more so than N2 sleep:***

Effects of sleep deprivation and NREM sleep on whole-brain respiration- and cardiac-driven brain pulsations were evaluated in 30sec epochs from well-rested (awake scans) and sleep deprived placebo (awake and NREM sleep scans) conditions. Whole-brain spectral power within individually tailored respiratory- and cardiac frequency bands were log transformed to mitigate skewness in the distribution before being analysed using a linear mixed model with the same random effect structure as LMM-*ii* and the same fixed effects with the addition of the base-10 logarithm of simultaneously recorded respiration or heart rates. $N$ = 20 (number of participants with data in each of the three conditions can be seen in Table S3).

Effects of sleep depth on spectral power within respiratory and cardiac frequency bands were evaluated in all 30sec epochs from the sleep deprived placebo condition classified as either wakefulness, N2 sleep or N3 sleep (*see* 'MREG data analysis'). Analyses were performed as described above, but with sleep depth (sleep-deprived awake vs N2 vs N3) included as an additive fixed effect together with respiration or heart rates (log transformed). $N$ = 19 (number of participants with data in each of the three conditions can be seen in Table S6).

P-values for testing fixed effects in both of the above linear mixed models were obtained using Wald tests. The expected log spectral power at the various conditions (e.g. sleep-deprived wakefulness) was computed from the mixed model estimates under mean respiration or heart rates of all 30-sec epochs included in the respective analyses – and used to illustrate the model fit (Fig 3E&G, Fig S4B-C and Fig 5B&D).

**(v) *EEG delta-power during MREG-imaging correlates with the MREG spectral power in respiration and cardiac frequency bands:***

The association between log spectral power in individually tailored respiration- and cardiac frequency bands and simultaneously recorded delta power ratios was evaluated in all 30sec epochs from well-rested and sleep deprived placebo scans, regardless of their visually scored vigilance and/or sleep states (*see* 'Quantitative analyses of EEG during MRI'*)*.

We performed a linear mixed model with the same random effect structure as LMM-*ii*, but with EEG delta power ratio and respiration or heart rates (log transformed) included as fixed effects. The reported p-value (denoted $p_{\text{adj}}$) is the one of the delta power ratio in the mixed model. The corresponding correlation coefficient ($r_{\text{adj}}$) was deduced from the mixed model estimates as described in *(i)* . For reference, Pearson correlation coefficients between delta power ratio and log spectral power in all included 30-sec epochs were evaluated (regardless of subject ID, respiration and heart rates, scan week and sleep opportunity) and denoted $r_{\text{raw}}$ (Fig 5F-G). $N$ = 20.

***(vi) NREM sleep primarily enhances spectral power in the respiration and cardiac frequency bands in grey and white matter:***

To explore whether the strength of brain oscillations differed between tissue types, we assessed spectral power within LFO, cardiac and respiratory frequency bands in grey matter, white matter and CSF. Data from well-rested (awake) and sleep deprived placebo (awake and NREM sleep) conditions were included in analysis and a linear mixed model with the same random effect structure as LMM-*ii* and the same fixed effects with the addition of tissue type (GM vs WM vs CSF) and respiration or heart rates (log transformed; only for resp./card. frequency bands) and the interaction between vigilance state and tissue type. P-values and estimates to illustrate the model fits (Fig 6C) were calculated as described in ***(iii)*** and ***(iv).*** *N* = 20 (number of participants with data in each of the three conditions can be seen in Tables S3 and S5).

***(vii) Adrenergic antagonism decreases spectral power in LFO and cardiac frequency bands***: Effects of carvedilol (randomised to sleep-deprived session 1 or sleep-deprived session 2) were assessed in data from sleep deprived placebo and sleep deprived carvedilol conditions.

Paired two-tailed Student's t-tests were used to compare blood pressure (Table S3) and serum norepinephrine levels (Fig S5), as well sleep characteristics during recovery sleep (REM latency, NREM total, REM total, Stage N2, stage N3; Table S4) between placebo and carvedilol conditions. For recovery sleep characteristics not normally distributed (TIB, TST, Sleep efficiency, Sleep latency, Stage N1; Table S4), Wilcoxon signed rank tests were used to compare the two conditions. *N* = 20. Linear mixed models were used to evaluate the effect of treatment on whole-brain log spectral power within LFO, respiration and cardiac frequency bands as well as on respiration and heart rates. Vigilance state (awake vs NREM sleep), treatment (placebo vs carvedilol) and respiration and heart rates (log transformed; only included for resp./card. frequency bands) and the interaction between vigilance state and treatment were included as additive fixed effects, while the random effect structure was similar to LMM-*ii*. P-values and estimates to illustrate the model fits (Fig 7) were calculated as described in ***(iii)*** and ***(iv).*** *N* = 20 (number of participants with data in each of the three conditions can be seen in Tables S3 and S5).

#### SUPPLEMENTARY REFERNCES

1. Klem, G.H., Lüders, H.O., Jasper, H.H., and Elger, C. (1999). The ten-twenty electrode system of the International Federation. The International Federation of Clinical Neurophysiology. Electroencephalogr Clin Neurophysiol Suppl *52*, 3–6.

2. Allen, P.J., Josephs, O., and Turner, R. (2000). A Method for Removing Imaging Artifact from Continuous EEG Recorded during Functional MRI. NeuroImage *12*, 230–239. https://doi.org/10.1006/nimg.2000.0599.

3. Niazy, R.K., Beckmann, C.F., Iannetti, G.D., Brady, J.M., and Smith, S.M. (2005). Removal of FMRI environment artifacts from EEG data using optimal basis sets. Neuroimage *28*, 720–737. https://doi.org/10.1016/j.neuroimage.2005.06.067.

4. Lipsitz, S.R., Leong, T., Ibrahim, J., and Lipshultz, S. (2001). A Partial Correlation Coefficient and Coefficient of Determination for Multivariate Normal Repeated Measures Data. Journal of the Royal Statistical Society: Series D (The Statistician) *50*, 87–95. https://doi.org/10.1111/1467-9884.00263.
